# Modeling the Impact of Active HPV Testing Intervention on Cervical Cancer Burden in India

**DOI:** 10.1101/2025.07.29.25332380

**Authors:** S Arvinth, Rashmi Tiwari, S Vishnu Tej, Piyush Sawarkar, Prerna Bhalla, Shekar Sivasubramanian, Ajit Rajwade, Ganesh Ramakrishnan, Nirav Bhatt

## Abstract

The global burden of mortality in women due to cervical cancer is relatively high in low and medium-income countries (LMIC), particularly India. The majority of cervical cancer cases are due to persistent and long-term infection with the Human papillomavirus (HPV). HPV testing plays a crucial role in identifying women with HPV infections at an early stage, before progression to cervical cancer, enabling timely intervention. This helps in reducing the cervical cancer burden in LMIC, in addition to vaccination and screening. However, there are no systematic modelling studies to investigate the role of HPV testing in cervical cancer burden. This work develops a mathematical model to investigate the impact of active HPV testing in reducing HPV infections and cervical cancer burden in India. By introducing an HPV infected compartment using HPV testing, our model captures the dynamics of known infected and unknown infected individuals, tracking infection progression, recovery from HPV infection, and cancer outcomes (cancerous cases, cancer mortality, cancer survival). The parameters in the proposed model are calibrated using the literature and epidemiological data specific to the Indian context. The simulation studies involving different testing population ages and HPV testing coverage rates are performed to understand the impact of active testing. The results from these studies indicate that higher testing rates can reduce the prevalence of cervical cancer and overall mortality. Furthermore, an optimal control approach with the active HPV testing coverage as a decision variable is employed to derive cost-effective strategies for minimizing treatment and HPV testing expenses. These findings highlight the necessity of appropriate resource allocation and policy interventions to curb the prevalence of HPV and mitigate the global burden of cervical cancer in India.

## 1. Introduction

Human Papillomavirus (HPV) infection is a global health concern, affecting millions of individuals worldwide, with significant implications for public health. Despite the availability of effective vaccines and screening technologies, the disease disproportionately affects low- and middle-income countries (LMICs), which account for nearly 85% of the global cervical cancer burden (Ginsburg et al. [10]). In India, HPV infection is a substantial health issue, with a diverse impact on different regions and demographics. The country bears a considerable burden of cervical cancer, where the disease is the second most frequent cancer among women, primarily attributed to high-risk HPV types. India’s vast and diverse population, coupled with variations in access to healthcare, contributes to the complexity of the prevalence and impact of HPV. Compared to the world, India faces a disproportionate burden of cervical cancer, with about 22 percent of global cases and deaths. The country’s HPV prevalence is double the global average. [27] has provided a comprehensive overview of incidence trends and mortality patterns based on National Cancer Registry Programme data.

HPV DNA testing is a genotype-based diagnostic test that can detect whether an individual is infected with HPV strains. The HPV DNA test is highly sensitive as it directly detects the presence of viral DNA, identifying women at risk before any cellular changes occur. It is a clinical process designed to detect the presence of high-risk HPV types (such as HPV-16 and HPV-18) that are associated with cervical cancer and other malignancies. While conventional cancer screening methods, such as cytology-based Pap smears, have played a role in early detection, they often fail to detect precancerous lesions in their earliest stages. HPV testing, on the other hand, is a more effective strategy because it detects high-risk HPV infections before cellular abnormalities develop, enabling early intervention and significantly reducing the risk of cervical cancer progression. Several studies have demonstrated the effectiveness of HPV-based screening compared to traditional cytology-based screening. Cox et al. [6] evaluated the impact of primary HPV testing versus 3-yearly cytology-based screening in New Zealand and found that HPV testing resulted in a greater reduction in cervical cancer incidence. Similarly, Brisson et al. [3] conducted a comparative analysis across LMICs, demonstrating that HPV testing, when combined with vaccination programs, accelerates the elimination of cervical cancer far more effectively than cytology alone. These findings highlight the superior sensitivity of HPV testing, allowing for earlier detection and treatment, thus preventing disease progression more efficiently than traditional screening methods. Arbyn et al. [1] also emphasized that HPV testing has superior sensitivity compared to cytology, allowing for earlier detection of precancerous changes and reducing cervical cancer incidence. Screening programs are intended to identify people at risk and intervene early to prevent cancer progression. Srinath et al. [30] systematically examined the barriers to HPV testing in India, identifying critical accessibility challenges and proposing solutions to increase screening uptake. HPV testing is a cornerstone of cervical cancer prevention. Poor Screening programs, along with socio-economic and health disparities, result in late diagnoses and thus poor outcomes, adding to India’s cervical cancer burden. Widespread implementation of Screening programs, particularly in Low- and Middle-income countries (LMICs) like India, could go a long way in reducing the incidence and mortality of cervical cancer, reducing cancer burden, and improving health outcomes for women, achieving WHO’s [34] target for cervical cancer elimination through early diagnosis and timely treatment.

Alternative screening approaches, such as HPV self-sampling and Visual Inspection with Acetic acid (VIA), have also been explored as cost-effective solutions to improve screening accessibility in underserved regions (Sauvaget et al. [28]). Sankaranarayanan et al. [25] studied HPV testing for cervical cancer in rural India in 2009 and conducted a clinical trial to measure the effect of a single round of screening by HPV DNA testing, cytologic testing, or visual inspection of the cervix with acetic acid (VIA) on the incidence of cervical cancer and the associated rates of death in a rural region in India. Additionally, HPV detection assays using urine, blood, or oral specimens have garnered growing attention in recent years due to their non-invasive nature and relative ease of self-collection Poljak et al. [23]. A meta-analysis study reported that the first void fraction of urine harbors significant concentrations of HPV DNA required for detecting genotype and genotype-specific viral load, thereby making it an effective specimen for HPV detection (Pathak et al. [21], Van Keer et al. [32]). Moreover, urine specimens can also aid in detecting anti-HPV antibody levels to monitor the immune status post HPV vaccination (Pattyn et al. [22]). Similarly, blood also contains detectable bio fragments, such as circulating HPV DNA (cHPV DNA) and/or circulating tumor DNA (ctDNA), collectively known as “ liquid biopsies” (Balachandra et al. [2]). The presence of such cell-free DNA serves as a potential biomarker indicative of underlying neoplastic processes. For instance, a meta-analysis published in 2020 evaluated the diagnostic accuracy of cHPV DNA for the invasive cervical cancer detection (Gu et al. [11]). The pooled sensitivity was reported to be 27% (95% CI: 24-30). Despite the well-established benefits of HPV testing, challenges remain in determining the most effective and resource-efficient screening strategies for large-scale implementation. The optimal frequency of screening, the target population for screening interventions, and the impact of integrating HPV vaccination with screening efforts are complex public health questions that require a systematic approach to evaluation. Given the diversity in healthcare infrastructure, population demographics, and disease burden across different regions, predictive tools are essential to guide policy decisions.

Mathematical modeling can carve out the complex dynamics of epidemiological processes from population and individual levels. By simulating HPV transmission, disease progression to cervical cancer, and the effects of interventions, models can provide data-driven insights into the potential outcomes of different screening policies. These models help policy and decision-makers to make informed decisions by considering the impact of many factors in scenarios otherwise marked by gaps and uncertainties. In particular, compartmental models allow researchers to track population dynamics across different disease states, providing a structured framework to assess the progression and regression of the infectious stages. Several authors have worked on evaluating the impact of HPV vaccination in the context of primary HPV testing and highlighted the effects of these interventions in reducing cervical cancer burden by using compartmental models. For instance, [16] and [35] developed mathematical models to assess how vaccination can complement primary screening programs in preventing high-risk HPV infections and associated disease progression. Similarly, [18] investigated the combined impact of HPV vaccination and screening strategies in previously unscreened populations. Similarly, Lee and Tameru [15] has developed a compartmental model of HPV in the United States and its impact on cervical cancer. Rajan et al. [24] has worked on developing a compartmental model for HPV in the Indian context. Gurmu and Koya [12] studied the role of testing as an intervention strategy in reducing the transmission of the disease. Work by Myers et al. [20] has worked on a Markov model for the HPV Infection and cervical carcinogenesis.

The primary objective of this research work is to develop a mathematical model of the Indian disease scenario to understand and forecast the dynamics of HPV and cervical cancer prevalence in India under the active HPV testing. Firstly, the model is formulated to include the Indian population dynamics over the next 75 years. Given no nationwide HPV testing programs in India and the lack of reliable prevalence data, the study relies on meaningful assumptions and predictions to estimate HPV disease burden for the model calibration. The model is calibrated to align with existing trends in disease and cancer prevalence by using the existing literature and epidemiological data for capturing the complex epidemiological landscape of India for meaningful forecasting. The calibrated model will evaluate the impact of active HPV testing on the reduction of cancer burden for different age cohorts of the female population. Finally, the optimal control problem is formulated to develop a cost-effective HPV testing to ensure the best possible outcomes for reducing cervical cancer burden in India.

The rest of this paper is structured as follows. The model with the HPV testing as a policy variable is formulated in Section 2. The model parameter calibration is performed in Section 2.2, using the existing literature values and model fitting on the epidemiological time-series data. Then, Section 2.4 formulates an optimal control problem to achieve cost-effective HPV testing coverage. Section 3 provides the results of model parameter estimation using the epidemiological data, sensitivity analysis, forecasting simulations studies for India and four major states, and the cost-effective interventions. Furthermore, the modeling results and role of HPV testing in reducing cervical cancer burden are discussed in Section 4. The findings from this study are concluded in Section 5.

## 2. Methodology

### 2.1. Model Formulation

The HPV Testing compartmental model proposed in this work consists of nine compartments involving susceptible population, infected population, cancerous population, and population associated with mortality, cervical cancer, and HPV recovery as shown in Figure 1. In this work, only the female population is considered. All females enter the model at a chosen age (for example, 18) and move between compartments in continuous time. Natural mortality (*μ*) is considered in every state. Throughout this section *N* (*t*) denotes the total simulated female population: *N* (*t*) = *S*_Target_ + *S*_Rest_ + *S*_Reinfec_ + *H*_*T*_ + *I* + *C* + *R*_*H*_ + *C*_*S*_ + *C*_*D*_. The time-varying inflow and outflow terms Λ_*j*_(*t*)*p*_*j*_ and *P*_*j*_(*t*),(*j* ∈ Target, Rest) are fitted and described in Appendix A for modelling the Indian population dynamics.

**Figure 1:**
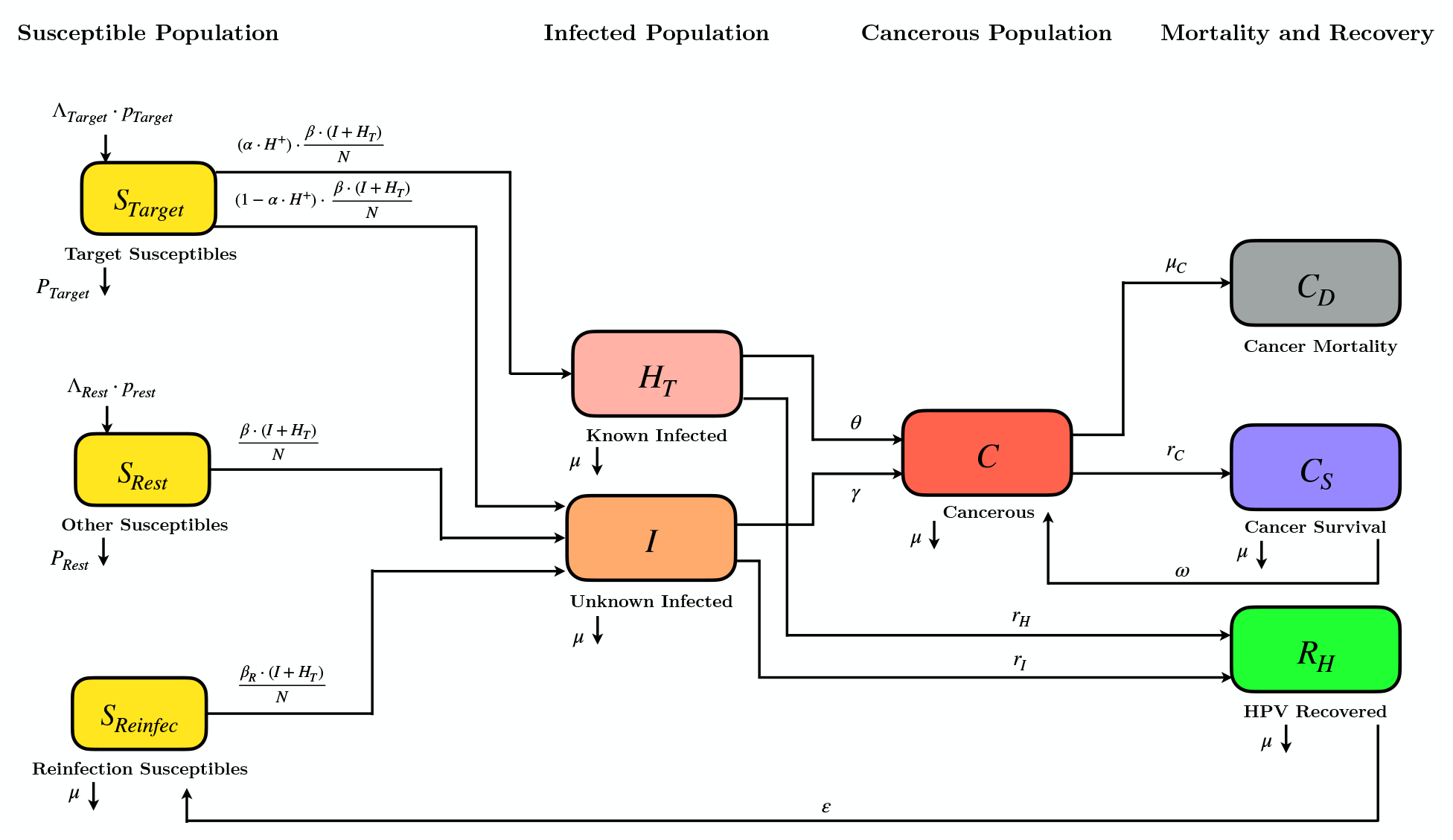
Compartmental Model for HPV and Cervical Cancer Dynamics with Age-Specific HPV Testing

#### 2.1.1. Susceptible population

The susceptible population consists of three subpopulations as follows:

1. Target Population for HPV Testing (denoted as *S*_Target_)
2. Rest of the susceptible female population outside the HPV Testing Target (denoted as *S*_Rest_)
3. Infected females who lost their immunity and are susceptible to reinfection (denoted as *S*_Reinfec_).

Susceptible target population being modeled as a separate compartment enables us to isolate a target age group for HPV testing intervention. For example, the entire female population aged 18 to 85 has been modeled, but if we would like to investigate the impact of the testing program only for females aged between 30 and 60. Then, these females are considered as the target population *S*_Target_ while the remaining females (18-29 and 60-85 age groups) do not come under this current intervention policy and fall into the *S*_Rest_ compartment. In addition to these two cohorts, the females who lost their immunity to the HPV infections (Compartment *R*_*H*_ in Fig. 1) after starting the HPV testing program are modeled as *S*_Reinfec_.

##### (i) Target susceptible Females S_Target_

This compartment holds females in the *policy–chosen age band* [*a*_min_, *a*_max_] (Equation 13 in Appendix A) who are currently eligible for organised HPV testing. This compartment allows for applying an age–specific testing coverage *α*(*t*) ∈ [0, 1]. *S*_Target_ has an incoming inflow of females from the target age band as a function of time, Φ_Target_(*t*) = Λ_Target_(*t*)⋅*p*_Target_, where Λ_Target_(*t*) is a Step-wise constant population function for the target age band which has been scaled with fitted scaling factors *p*_Target_ (refer to Equations 14 and 15, in Appendix A) and an outflow of females also as a function of time, Ψ_Target_(*t*) = *P*_Target_(*t*) (refer to Equation 14). Both the inflows and outflows are calibrated for *S*_Target_ to match the population growth dynamics of Indian females in the target age band. A portion of females from the *S*_Target_ compartment can get HPV tested, with *HPV Testing Coverage Rate, α*, with the outcome either as being tested *Positive* or *Negative* for HPV. Here, *α* is multiplied with *HPV Testing Success Rate H* ^+^ to account only for the True-Positive HPV tested females (*α* ⋅ *H* ^+^). Similarly, (1 − *α* ⋅ *H* ^+^) accounts for the people who are not part of this HPV testing coverage. The HPV positive females under the HPV testing program (*α* ⋅ *H* ^+^) can get infected with the *infection transmission factor* of 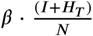 and move to the known-infected compartment, *H*_*T*_. Females who were not part of this HPV testing coverage (1− *α* ⋅ *H* ^+^) can get infected with the same infection transmission factor of 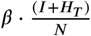 but move to the unknown infected compartment *I*.

##### (ii) Rest susceptible Females S_Rest_

This compartment holds females aged 18 to 85 who lie outside the target age band for testing, and remain susceptible. *S*_Rest_ has an incoming inflow of women from the rest age band as a function of time, Φ_Rest_(*t*) = Λ_Rest_(*t*) ⋅ *p*_Rest_, where Λ_Rest_(*t*) is a step-wise constant population function for the rest age band which has been scaled with fitted scaling factors *p*_Rest_ (refer to Equations 14 and 15, in Appendix A) and an outflow of females also as a function of time, Ψ_Rest_(*t*) = *P*_Rest_(*t*) (refer to Equation 14). The inflows and outflows have been calibrated for *S*_Rest_ to match the population growth dynamics of Indian females who are not in the target age band. Females from the *S*_Rest_ compartment can get infected with HPV with the same *infection transmission factor* of 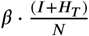 and moved to the unknown-infected compartment, *I*.

##### (iii) Reinfection susceptible S_Reinfec_

HPV recovered females from *R*_*H*_ gradually lose type–specific immunity at rate *ε* and become vulnerable for re-infection, hence, moving to *S*_Reinfec_. From *S*_Reinfec_, females can get re-infected with the *re-infection transmission factor* 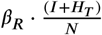 and move to unknown infected compartment, *I*.

The model equations for the susceptible popu^*N*^lation are as follows:

###### Susceptible Population

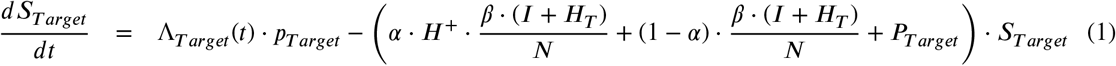

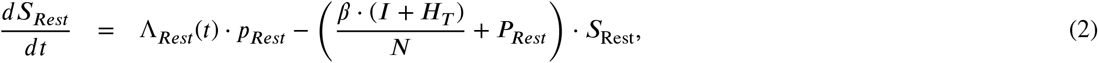

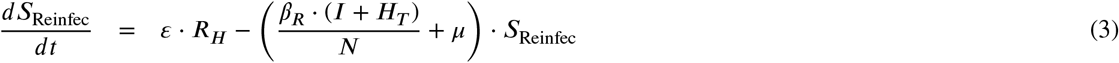

#### 2.1.2. Infected population

The infected populations are distinguished according to testing status into:

1. Unknown Infected (Not HPV tested and unaware of their infectious state, denoted as *I*)
2. Known Infected (HPV tested, Infected and aware of their infectious state, denoted as *H*_*T*_)

These infected populations have two outcomes: (i) their condition can worsen and they can progress to the Cervical Cancer compartment, *C*, or (ii) they can recover from the HPV infection and move to the HPV recovered compartment, *R*_*H*_.

##### (iv) Unknown infected females I

This compartment aggregates (a) target females who are not tested, (b) all primary infections in *S*_Rest_, and (c) reinfections from *S*_Reinfec_. The unaware-infected females can progress to cervical cancer compartment *C*, with cancer progression rate *γ*, or recover naturally and move to HPV recovered compartment *R*_*H*_, with recovery rate *r*_*I*_.

##### (v) Known infected females H_T_

Females tested for HPV *and* identified positive with the HPV enter *H*_*T*_ as mentioned in Subsection 2.1.1. It is hypothesized that earlier awareness (leads to behavioural change) and possible treatment is expected to slow the progression to cancer *θ < γ* (slower cancer progression rate of Known Infected *H*_*T*_ relative to Unknown Infected *I*) and speed viral clearance leading to faster recovery *r*_*H*_ *> r*_*I*_, i.e., faster HPV recovery rate of Known-infected *H*_*T*_ relative to Unknown Infected *I*.

The model equations for the Infected population are as follows:

###### Infected Population

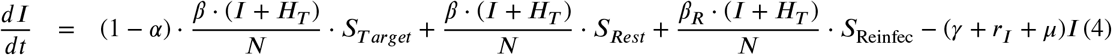

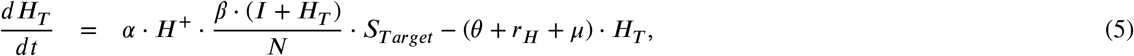

#### 2.1.3. Cancer related Population

The cancer-related population consists of three subpopulations as follows:

1. Cervical Cancer (HPV infected females who have progressed to Cervical Cancer, denoted as *C*)
2. Cervical Cancer Survivors (denoted by *C*_*S*_)
3. Cancer Mortality (Women who have expired due to Cervical Cancer, denoted by *C*_*D*_).

##### (vi) Cancer C

Incident cervical cancer cases arise from both known Infected *H*_*T*_ and unknown Infected *I* compartments as seen in Subsection 2.1.2 and relapses from cancer survival compartment *C*_*S*_. Cervical cancer patients from compartment *C* can either expire and move to Cancer Mortality Compartment *C*_*D*_, or survive and move to the Cancer Survival Compartment *C*_*S*_.

##### (vii) Cancer Survival C_S_

Survivors from Cervical Cancer compartment *C* enter Cancer Survival Compartment *C*_*S*_ with the survival rate *r*_*C*_, improve after successful therapy, but face an elevated relapse risk of moving back to *C* compartment with the Cervical Cancer relapse rate *ω*.

##### (viii) Cancer Mortality C_D_

Mortality from cervical cancer compartment *C* accumulates as in the Cancer Mortality compartment *C*_*D*_ with the Cervical Cancer mortality rate *μ*_*C*_.

The model equations for the cancerous population are as follows:

###### Cancerous Population

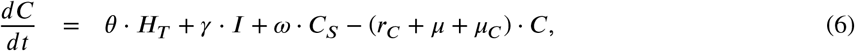

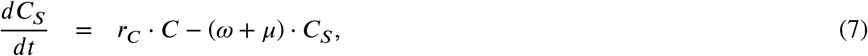

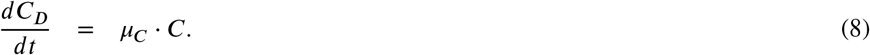

#### 2.1.4. Recovered Population

##### (ix) HPV Recovered R_H_

Natural or treatment–assisted clearance from both known Infected *H*_*T*_ and unknown Infected *I* compartments confers temporary immunity, which can wane at a rate of *ε*, causing HPV recovered females to be susceptible to reinfection by moving them to *S*_*Reinfec*_.

The model equation for the recovered population is as follows:

###### Recovered

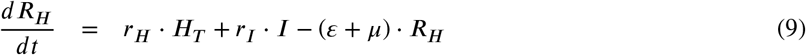

### 2.2. Model Parameter Calibration

All the compartments, mainly, *S*_Target, Rest_, *I, C*, and *C*_*D*_ of the mathematical model should follow the existing population growth and epidemiological trend, for reliable forecasting and assessment of the impact of interventions. The parameters for the model are calibrated by three distinct approaches. First, parameters such as *μ, ε, r*_*I*_, *r*_*C*_ and *ω* are determined based on values sourced from established literature. Second, parameters Λ_Target_, Λ_Rest_, *p*_Target_, *p*_Rest_, *P*_Target_,*P*_Rest_,*β, β*_*R*_, *γ* and *μ*_*C*_ are estimated through fitting using population growth, mortality and epidemiological or data. Lastly, parameters *θ* and *r*_*H*_ are assumed within the framework of this study to align with the specific objectives of the model calibration. The list of parameters and their corresponding meaning used in the model, along with their value is summarized in Table 1.

**Table 1.** List of Parameters for HPV Testing Model.

| Parameter | Definition | Final Value | Source |
| --- | --- | --- | --- |
| $H^+$ | HPV Testing Success Rate | 0.95 | From literature [19] |
| $\beta$ | HPV Infection Transmission Rate | 0.5433 | Estimated from data |
| $\beta_R$ | Re-infection Transmission Rate of Previously HPV Infected Population | 0.4932 | Estimated from data |
| $\alpha$ | HPV Testing Coverage Rate | Control Variable | - |
| $\gamma$ | Progression Rate of Unknown HPV Infected to Cancerous | 0.0012834 | Estimated from data |
| $\theta$ | Progression Rate of Known HPV Infected to Cancerous Stage | $\theta \sim \mathcal{N}\left(\frac{\gamma}{2}, \left(\frac{\gamma}{10}\right)^2\right)$ | Modeled/ Assumed |
| $r_I$ | Recovery Rate of Unknown HPV Infected | 0.45 | From literature [17] |
| $r_H$ | Recovery Rate of Known HPV Infected | $0.45 \leq r_H \leq 1 \text{ year}^{-1}$ | Modeled / Assumed |
| $r_C$ | Survival Rate of Cancerous Population | 0.1034 | From literature [26] |
| $\mu$ | Natural Mortality Rate of Population | 0.01389 | From literature [31] |
| $\mu_C$ | Mortality Rate of Cancerous Population | 0.0008526 | Estimated from data |
| $\epsilon$ | Rate of losing immunity and becoming infected again | 0.35 | From literature [33] |
| $\omega$ | Cancer Relapse Rate | 0.162 | From literature [14] |

### 2.3. Parameters computed or obtained from literature

#### HPV Testing Success Rate, H ^+^

We adopt the value of *H* ^+^ = 0.95 to represent the HPV testing sensitivity, based on a pooled estimate from a systematic review and meta-analysis conducted by Mustafa et al. [19]. This review synthesized results from five studies found that the HPV test had a sensitivity of 0.95 (95% CI: 0.84–0.98). The sensitivity of different HPV tests is discussed in Appendix B.

#### Unknown HPV Infected Recovery Rate, *r*_*I*_

Johns Hopkins Medicine [17] states that, for 90% of women with HPV, the condition will clear up on its own within two years. From this, *r*_*I*_ is calculated as,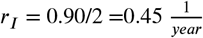.

#### Cervical Cancer Survival Rate, *r*_*C*_

From the study on Survival of patients with cervical cancer in India by Sathishkumar et al. [26], the overall pooled data of cervical cancer free five-year survival was 51.7%. From this, *r*_*C*_ is calculated as: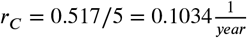.

#### Natural Morality Rate, μ

The Natural Mortality rate has been calculated from the UN Data [31], where the average age of mortality/lifetime of women in India was found out to be 72. Hence,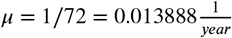.

#### Rate of Losing Immunity, ε

Wentzensen et al. [33] studied naturally acquired immunity to HPV infections in a subgroup. The combined odds ratio for the HPV16 and HPV18, the virus-like particles (VLP) based ELISA negativity was 0.65 (95% CI, 0.46–0.93). Hence, the odds of them being infected are *ε* = 1 − 0.65 = 0.35.

#### Rate of Cervical Cancer Relapse, ω

The study by Kumar et al. [14] on the overall survival in patients with atypical and typical recurrences of cervical cancer showed that a total of 225 (16.2%) out of 1719 patients had documented cervical cancer recurrences. Using the study data, the cervical cancer relapse rate is taken as *ω* = 0.162.

#### Parameter Estimation

The parameters *β, β*_*R*_, *γ*, and *μ*_*C*_ of the compartmental model are estimated by formulating a least-squares problem. The optimization problem minimizes the sum of squares between the model predictions and the actual data for the infectious compartment (*I*), the cervical cancer compartment (*C*), and the cancer mortality compartment *C*_*D*_, with the assumption of the remaining parameters taken from the literature as mentioned earlier.

#### Infection Data

HPV Information Center [4], has a collective data of numerous studies conducted all over India, on varying cohort sizes, age groups, etc. The studies for the highly infectious variant *HPV 16/18*, are mentioned in the Table 4 in the Appendix section. Since no nationwide HPV testing program was implemented, the HPV infection burden data are not available. The study data in [4] for India in Table 4 are used to compute the HPV prevalence. The mean of the *Prevalence Percentages* is calculated, mean prevalence % = 5.2%. It is assumed that 5.2% of the susceptible female population carries the highly infectious variant HPV 16/18.

#### Cervical Cancer Data

The Cervical Cancer prevalence from the year of 1990-2021 is available in the Institute for Health Metrics and Evaluation, Global Disease Burden (GBD) [13].

#### Cervical Cancer Mortality Data

The Cervical Cancer Mortality from the year of 1990-2021 is also available in the Institute for Health Metrics and Evaluation, Global Disease Burden (GBD) [13].

With the HPV-testing coverage fixed at *α*(*t*) ≡ 0, the known infectious compartment *H*_*T*_ disappears and the full model collapses to an **SICR** core. The surveillance data (*I*_actual,*t*_, *C* _actual,*t*_, *C*_actual,*t*_) From 1990–2021 are used as the actual data. The weighted least-square objective function with parameter vector **q** = (*β, β*_*R*_, *γ, μ*_*C*_)^⊤^ is

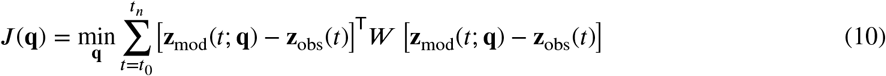

where **z**_obs_(*t*) = (*I*_actual,*t*_, *C*_actual,*t*_, *C*_*D* actual,*t*_)^⊤^ and **z**_mod_(*t*; **q**) = (*I*_model,*t*_, *C*_model,*t*_, *C*_*D* model,*t*_)^⊤^ predicted using Equations (1)-(9). The feasible set for the parameter values is given as {Ω = 0.2 ≤ *β, β*_*R*_ ≤ 0.8, 10^−4^ ≤ *γ* ≤ 2 × 10^−2^,0 ≤ *μ*_*C*_ ≤ 1}. The weighted least-squares problem is minimized under the box constraints provided by the feasible set.

#### Modeling of θ

The parameter *θ*, representing the progression of the screened infected population to cervical cancer, is a critical factor in determining the effectiveness of the screening program. It is defined within the bounds:

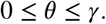

These bounds reflect the extent to which screening can mitigate the progression of an infected individual to cervical cancer, depending on the timing of the screening. The lower bound (*θ* = 0) corresponds to an ideal scenario where an individual is screened immediately after infection, maximizing the likelihood of preventing progression to cancer. Conversely, the upper bound (*θ* = *γ*) represents the worst-case scenario where an individual has been infected for an extended period before screening, resulting in no discernible benefit from screening compared to an unscreened individual. Hence, *θ* has been modeled as a normal distribution with standard deviation *γ*/10 and mean *γ*/2.

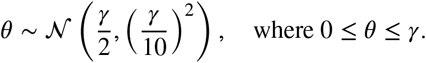

#### HPV Known, Infected Recovery Rate, r_H_

There is no solid information available on how HPV testing of a patient decreases their recovery time, relative to someone who is untested and infected. Hence, we assume that 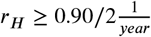.

### 2.4. Cost effective HPV testing Intervention using Optimal Control Formulation

In order to identify the cost-effective HPV testing (testing rate *α*(*t*)) over the campaigning years, a control problem that minimizes the total cost, HPV testing and cervical cancer cost, is formulated. The central idea of this control problem is to find the cost-effectiveness of the HPV testing, if any, with respect to the treatment cost. The optimal control problem with *α*(*t*), HPV testing rate, is formulated with the HPV dynamics models involving a nine-dimensional state system in Section 2, Eqs. (1)–(8). It is assumed that policymakers can manipulate directly the age-specific HPV-testing coverage rate, denoted by the measurable control function *α*(*t*) ∈ [0, 1]. To find the time-path *α*^*^(*t*) that minimises the cumulative economic burden of cervical cancer while accounting for programme costs, the following optimal control problem is solved

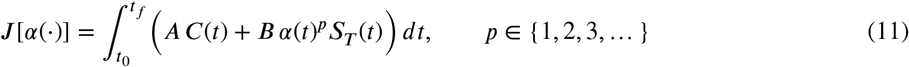

subject to the HPV dynamical equations Eqs. (1)–(8), 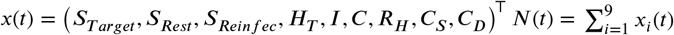 and *α* ∈ [0, 1]. where *A* represents the cost associated with the treatment of cervical cancer patients, and *B* denotes the cost per unit of HPV testing intervention. *p* ∈ {1, 2, 3, …} = Cost Power (1 = linear, 2 = quadratic …), is taken as *p* = 8. The parameter *α* is the HPV testing coverage rate, *C*(*t*) is the cancer population at time *t*, and *S*_Target_(*t*) is the susceptible women in the chosen target age band at time *t*. The optimal control problem in Equation (11) is solved using the indirect solution method by constructing the Hamiltonian as follows

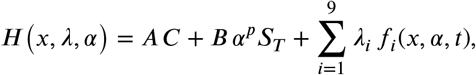

where *f*_*i*_ are the right–hand sides of (1)–(8) in the same order (*S*_*T*_, …, *C*_*D*_). The necessary conditions of optimality using Pontryagin’s principle are derived. The details of the optimality conditions and the numerical algorithm steps to solve the problem are provided in Appendix F.

## 3. Results

### 3.1. Calibrated Model using Parameter Estimation

The baseline calibration (*α*(*t*) ≡ 0) quantifies key transmission and progression parameters before any HPV testing intervention. Using the weighted least-squares criterion in Eq. (10), the four parameters **q** = (*β, β*_*R*_, *γ, μ*_*C*_)^⊤^ were fitted to the national series 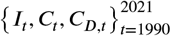. The remaining epidemiological constants were fixed at the values in Table 1, and the initial stocks *I*_1990_, *C*_1990_, *C*_*D*, 1990_ were set equal to the first observations. The simulation prediction is compared with the existing epidemiological scenario in Figure 2 compares simulated trajectories (solid lines) with observed data points (red markers) for the period 1990–2100. The fitted model adequately describes the behaviour of different compartments. Hence, the calibrated model is used for further analysis.

**Figure 2:**
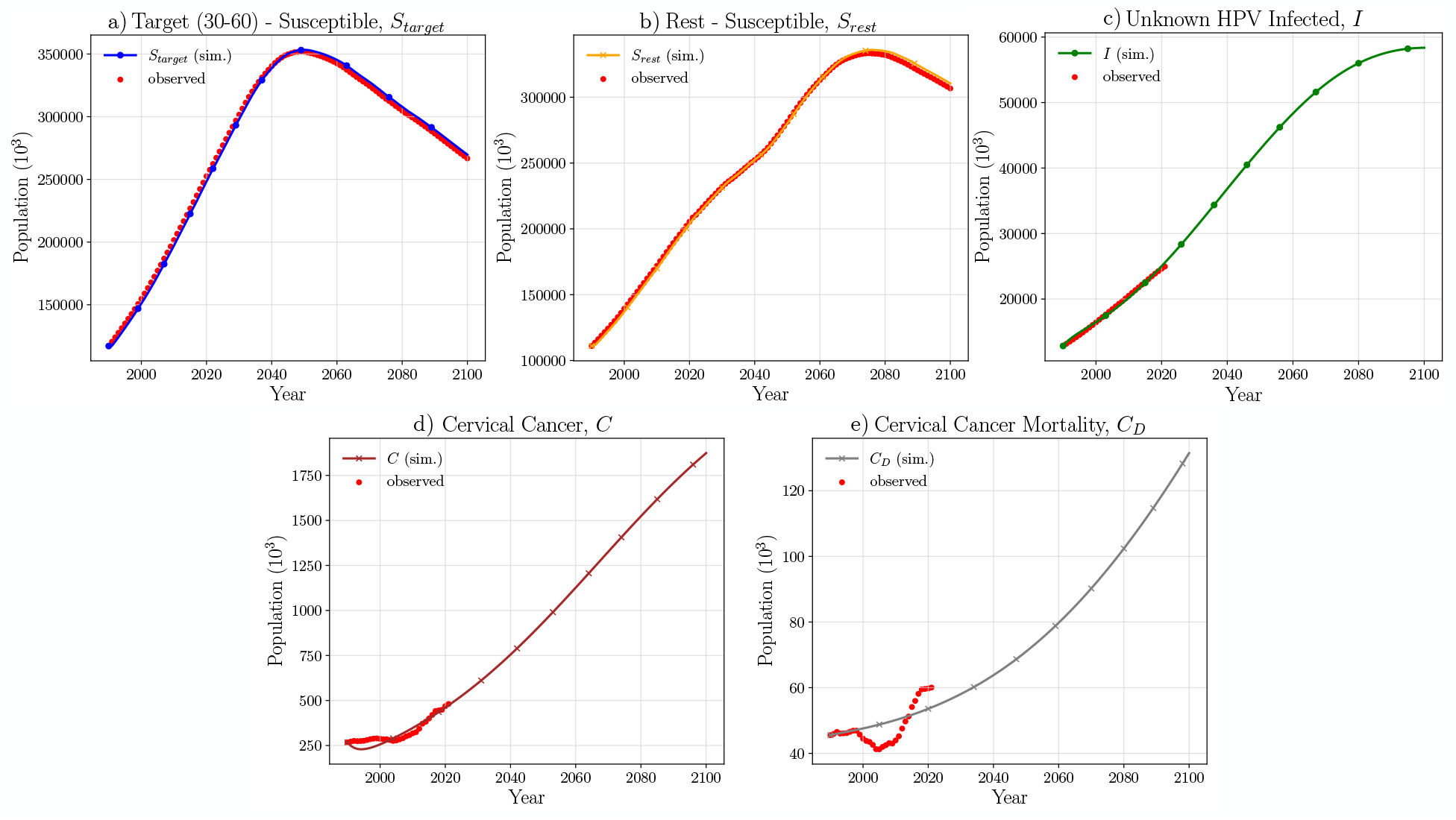
Comparison of Actual values vs. Simulation Prediction for compartments: a) *S*_Target_, b) *S*_Rest_, c) *I*, d) *C* and e) *C*_*D*_.

#### Sensitivity Analysis of Model Parameters

The model parameters in the final calibrated model are subjected to a sensitivity analysis for capturing the impact of various parameters on the cancer prevalence dynamics. The sensitivity analysis is performed by sampling a range of nine values with symmetrical variation around the original parameters in the calibrated model. The corresponding effects on the cancer incidence (*C*) were analyzed over a projected time period from 1990 to 2100. The parameters are varied across a predefined range (decided upon the confidence intervals of those respective parameters), reflecting potential variability and uncertainty in estimations. Figure 3 presents the results for all the model parameters. *H* ^+^ (HPV Testing Success Rate), *θ* (Cervical Cancer Progression Rate of Known HPV Infected Population), *μ*_*C*_ (Cervical Cancer Mortality rate) and *ω* (Relapse rate of Cervical Cancer Survivors) show very low sensitivity towards Cervical Cancer Compartment. Where as *β* (HPV infection transmission rate), *β*_*R*_ (HPV re-infection transmission rate), *γ* (Cervical Cancer progression rate of Unknown Infected Population), *r*_*I*_ (Natural Recovery rate of Unknown Infected population) and *ε* (waning immunity rate) are highly sensitive towards the Cervical Cancer Compartment in the model. For completeness, Appendix C derives an explicit closed-form expression for the *Basic Reproduction Number, ℛ*_0_, of the SICR core using the next-generation matrix method of van den Driessche and Watmough [9]. Appendix D then applies the normalised sensitivity-index formula to quantify how relative changes in each epidemiological parameter propagate into ℛ_0_.

**Figure 3:**
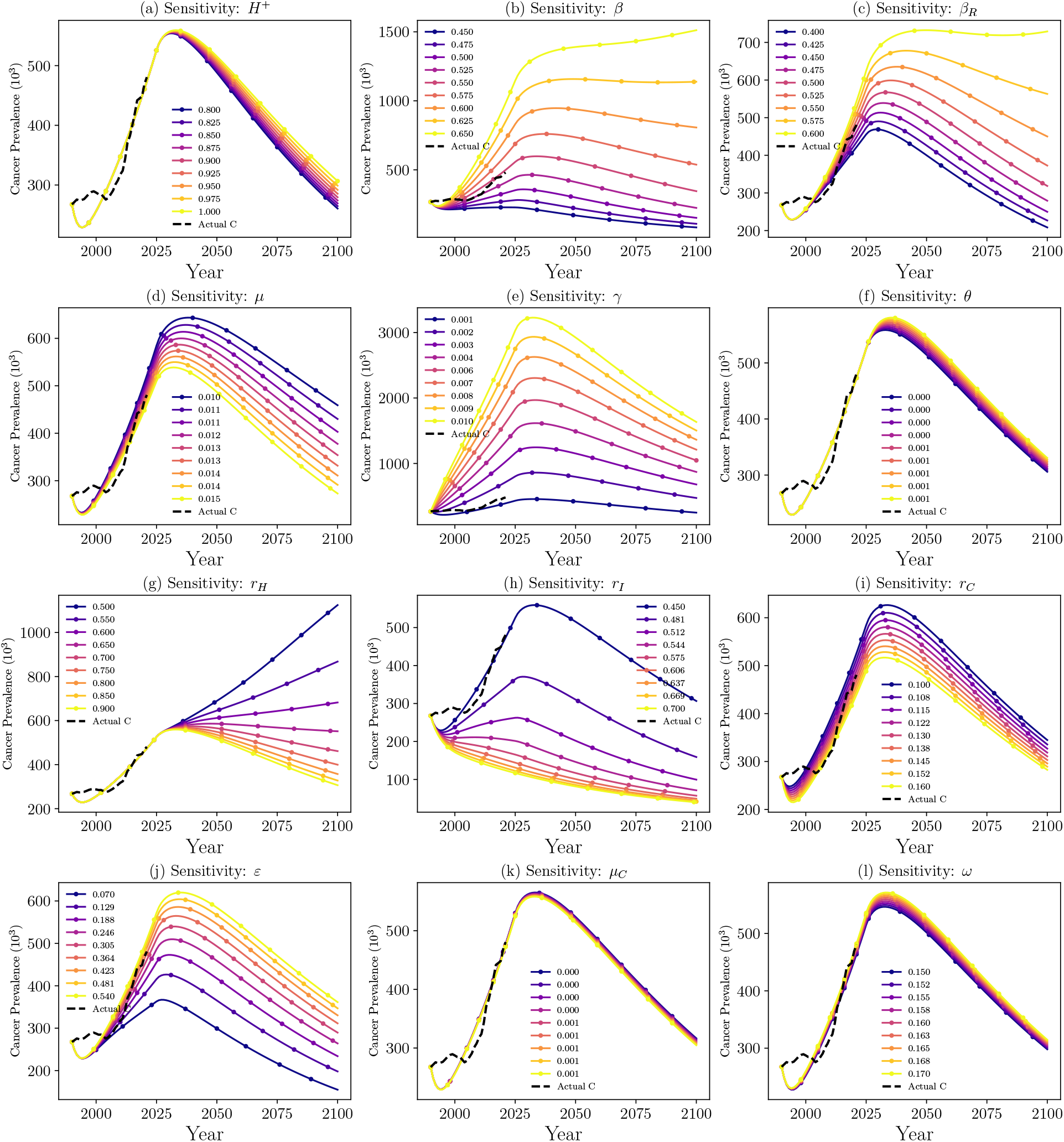
Sensitivity Analysis of Model Parameters (With 90% Testing Coverage)

### 3.2. Forecasting Simulation Studies

This section investigates the impact of different testing coverage rates (*α*(*t*)) and the different target age groups for HPV testing at constant coverage rates on HPV infection and cervical cancer cases through a set of forecasting simulations.

#### 3.2.1. Investigating Different Testing Coverage Rates, α

To comprehend the effect of testing intensity on all states of the model, a forward simulation is performed in which the HPV-testing coverage parameter *α* is varied over the set {0.00, 0.25, 0.50, 0.75, 0.90, 1.00}. Figure 4 shows the trends of all the compartments for the HPV Testing rates *α* = [0, 0.25, 0.50, 0.75, 0.90, 1.00]. All epidemiological parameters remain unchanged (Table 1).

**Figure 4:**
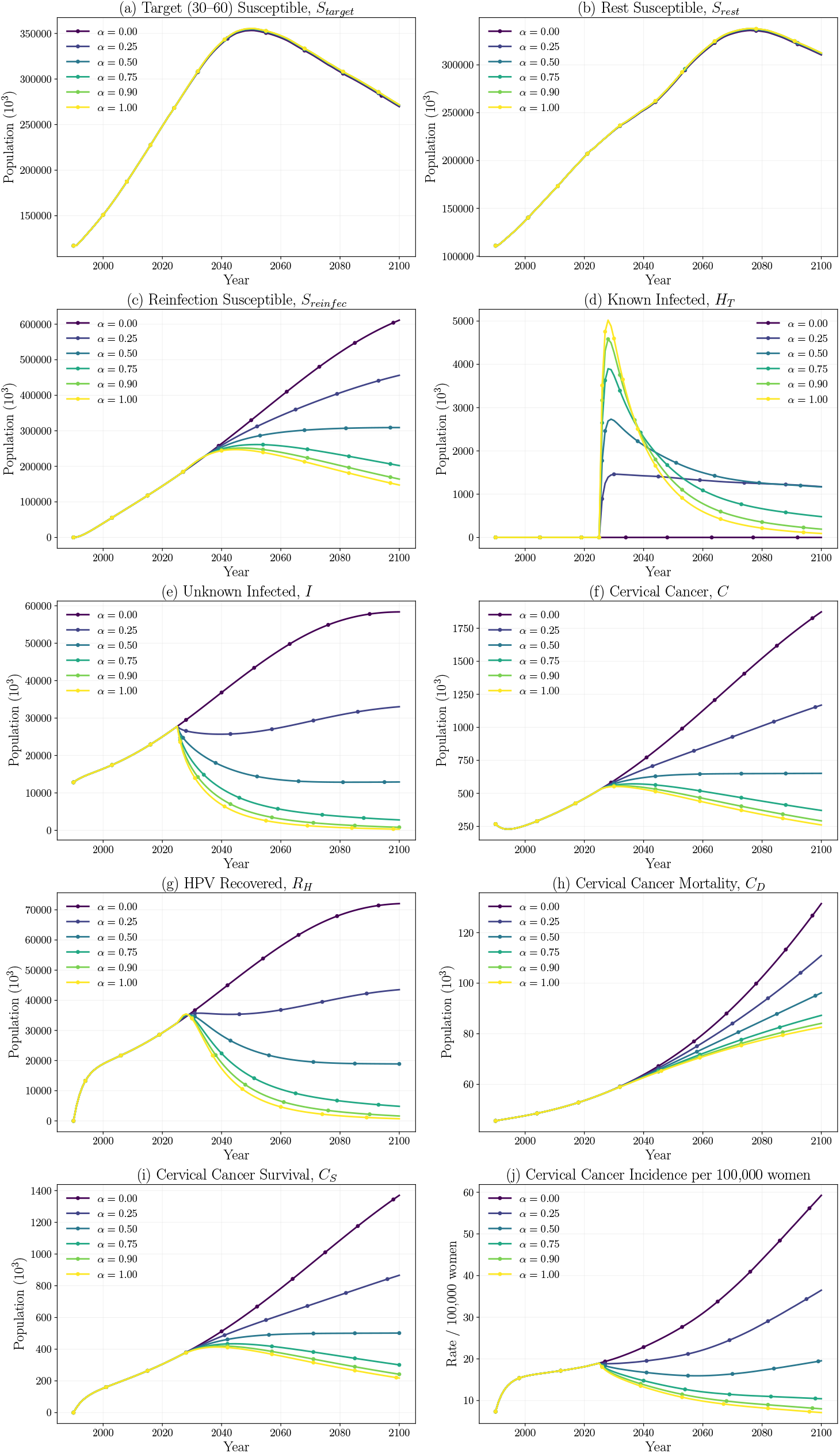
Effect of HPV Testing Rate *α*, on on nine state variables (a)-(i) and Cervical-Cancer Incidence per 100K women (j)

In the *HPV Testing Infected (H*_*T*_ ) compartment, higher *α* implies an increased number of females tested for HPV infection, thereby resulting in a steeper rise and stabilization of the *H*_*T*_ population. The *Infected Individuals I* [Figure 4(e)], and *Cancer Cases C* [Figure 4(f)] compartments exhibit a noticeable peak and subsequent decline over time. Moreover, with higher values of *α*, a rapid reduction in infected population, a steeper growth in *Recovered Populations R*_*H*_ [Figure 4(g)] and slower increase in *Cancer Deaths C*_*D*_ [Figure 4(h)] was observed. This observation indicates a positive impact of the increase in HPV testing rates on reducing the HPV infection and cervical cancer burden.

#### 3.2.2. Investigating Different Target Age Groups for HPV Testing

Figure 5 shows the trends of all the compartments for the different choices of targeted age bands of [18 − 70*y*, 30 − 60*y*, 30 − 85*y*, 25 − 65*y*, 25 − 35*y*] at the constant rate of HPV testing coverage of 65% (*α* = 0.65). These target bands were chosen after carefully reviewing HPV testing programs carried out in different countries ([7], [5]). It can be observed from Figure 5, we observe that expanding the target age band for HPV testing leads to consistently greater reductions in the burden of HPV infection and cervical cancer across all key compartments in the model. The number of *HPV-known infected* individuals *H*_*T*_ [Figure 5(d)] increases sharply for wider bands due to greater testing coverage. The *unkown infected population I* [Figure 5(e)] decreases most significantly for broader bands. The *cervical cancer* compartment *C* [Figure 5(f)] and associated *cancer mortality C*_*D*_ [Figure 5(h)] reduce the most under the 18–70y. The *recovered individuals* from HPV, *R*_*H*_ [Figure 5(g)] are higher for wider bands.

**Figure 5:**
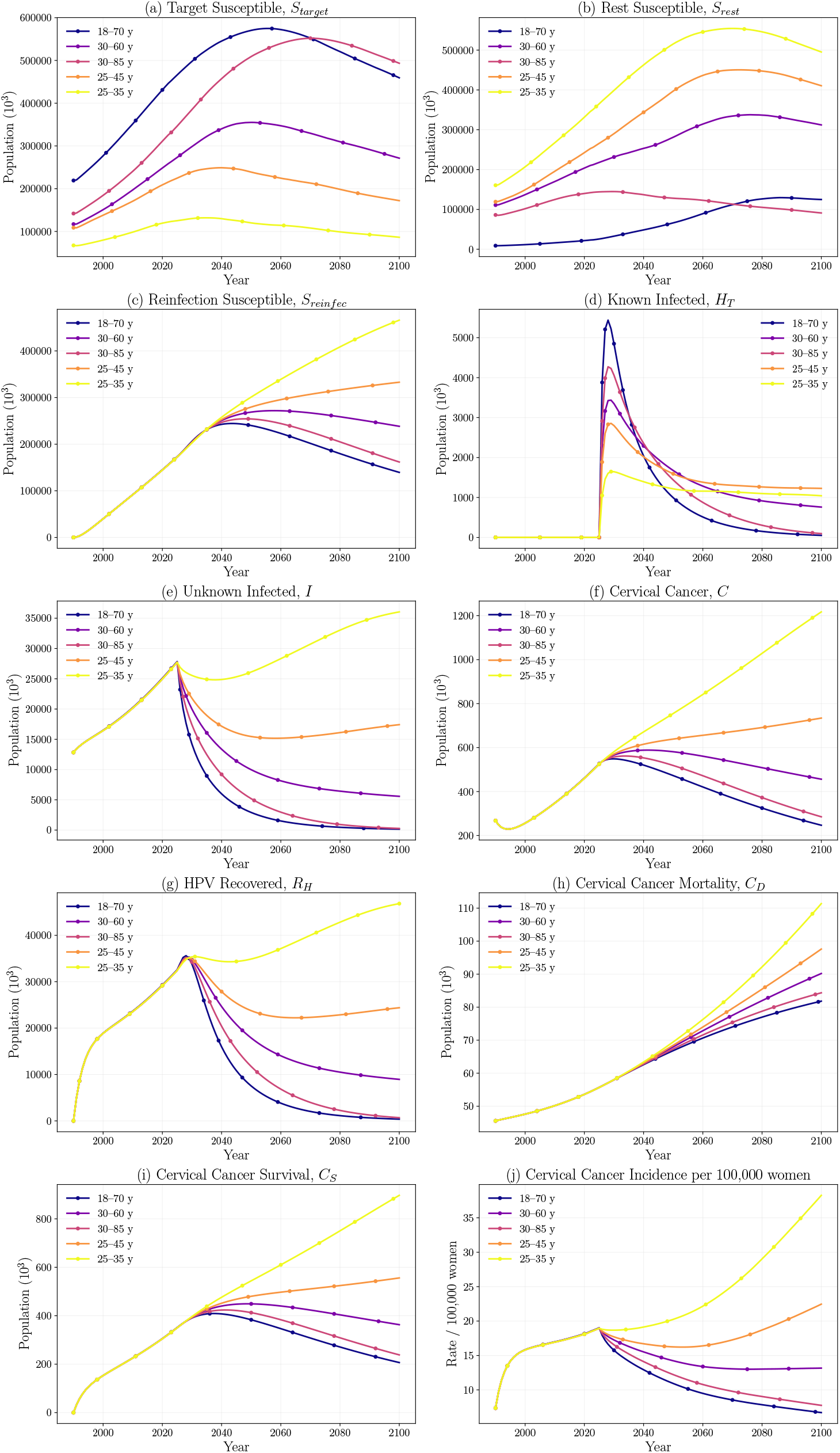
Effect of choosing different Target Age Groups for Testing, on all nine state variables (a)-(i) and on Cervical Cancer Incidence per 100K women (j) (With 65% Testing Coverage)

#### 3.2.3. State-wise Forecasting Simulation Study of HPV Testing Impact

To assess the state-level benefits of HPV testing, we simulated cervical cancer prevalence trajectories across four major Indian states, namely, West Bengal, Maharashtra, Tamil Nadu, and Uttar Pradesh, under two screening scenarios: Baseline (No Testing) where, *α*(*t*) ≡ 0 and HPV Testing (65% Coverage): where *α*(*t*) = 0.65 for *t* ≥ 2025 Each state-specific simulation incorporates demographic structure, calibrated transmission parameters (*β, β*_*R*_), cancer progression rates (*γ*), and cancer-specific mortality (*μ*_*C*_) fitted separately for each state. The model is simulated from 2011 to 2100 using the state-specific initial conditions for infections and cancer burden for forecasting purposes.

Figure 6 compares projected cervical cancer incidence per 100,000 women under the two scenarios: Baseline (No HPV Testing, 0% Coverage) and HPV Testing (65% Coverage) at three time points in the future: 2040, 2060, and 2080. For *West Bengal* [Figure 6(a)], the 65% HPV-testing coverage reduces incidence from ≈ 14 to 11 cases per 100 000 women in 2040 (23% reduction), from 16 to 10 cases in 2060 (39% reduction), and from 20 to 11 cases by 2080 (47% reduction). *Tamil Nadu* [Figure 6(c)] begins with the highest baseline incidence, and screening lowers the rate from 21 to 17 cases in 2040 (22% reduction), 24 to 15 cases in 2060 (39% reduction), and 29 to 15 cases by 2080, achieving a 48% reduction. Similar trends are observed for Maharashtra and Uttar Pradesh.

**Figure 6:**
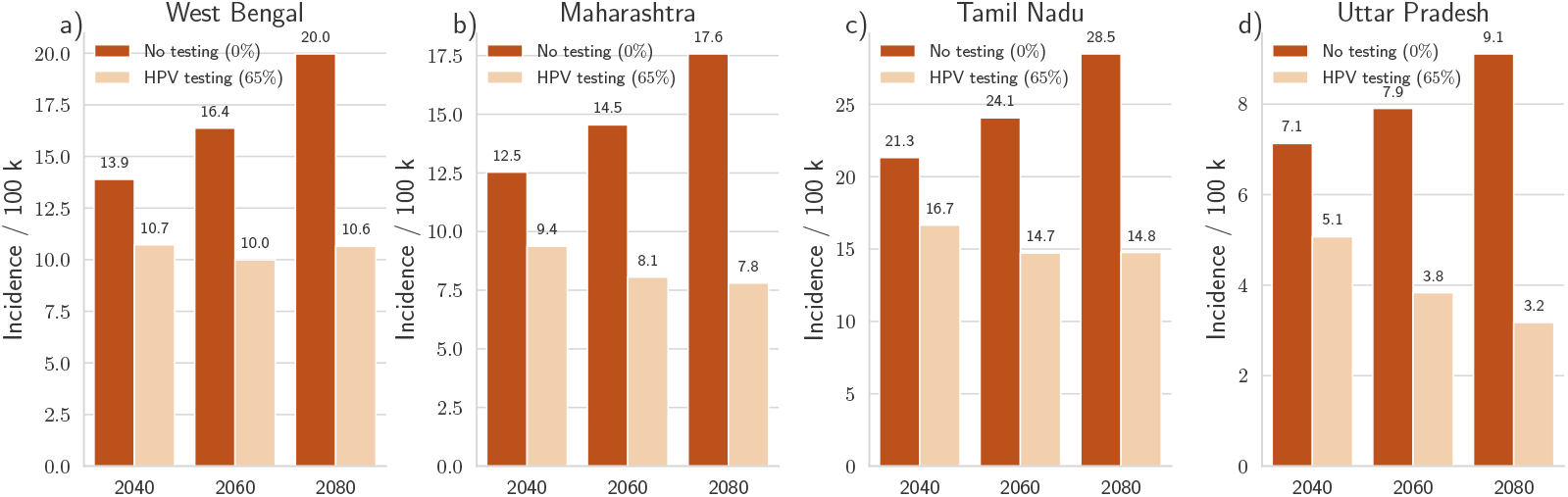
Projected Cervical Cancer Incidence per 100,000 women in four Indian states: a) West Bengal, b) Maharashtra, c) Tamil Nadu, d) Uttar Pradesh, under two screening scenarios: Baseline (0%) vs HPV Testing (65%). Incidence is shown for the years 2040, 2060, and 2080. Simulations use state-specific parameters and initial conditions calibrated to 2011 data.

### 3.3. Cost-effective HPV Testing using Optimal Control Formulation

The investigation of the optimal HPV testing coverage over time, *α*(*t*) is performed from 2025 to 2100 with the target age band of *30-60* after reviewing the screening programs compiled in Cuzick et al. [7] and Chrysostomou et al. [5]. The rationale for adapting a rounded conservative upper-bound for the per-patient Cervical Cancer treatment cost (refer to Equation 11) of *A* = 60, 000INR (≈ USD 720) has been decided based on the estimates from Singh et al. [29], and HPV DNA testing per female of (refer to Equation 11) of *B* = 3, 000 INR (≈ USD 36), both of which can be found in Appendix G which provides a conservative yet empirically grounded estimate for cost-impact calculations.

The optimal trajectories of *S*_target_, *S*_rest_, *S*_reinfec_, *H*_*T*_, *I, C, R*_*H*_, *C*_*S*_ and *C*_*D*_ compartments are plotted in Figure 8 [(a)-(j)]. The corresponding *Optimal HPV Testing Rate, α*^*^(*t*), necessary for obtaining these optimal trajectories for the nine state variables can be seen in Figure 7(a). The optimal HPV testing coverage *α*^*^(*t*) begins at 60% coverage in 2025 and gradually decreases over the following decades. Under this optimal testing strategy, the *HPV known infected* compartment, *H*_*T*_ [Figure 8(d)] shows an early peak increase followed by a sharp decline, indicating aggressive detection followed by containment. The *unknown infected population, I* [Figure 8(e)] steadily declines after the control is introduced. The *cervical cancer* compartment *C* [Figure 8(f)], *cancer mortality, C*_*D*_ [Figure 8(h)], and *survival, C*_*S*_ [Figure 8(i)] reflect significant improvement compared to uncontrolled trajectories. Specifically, *C* and *C*_*D*_ decline continuously, showing the long-term benefits of sustained, cost-optimized HPV testing. The increase in *C*_*S*_ [Figure 8(i)] illustrates improved outcomes due to earlier detection and treatment.

**Figure 7:**
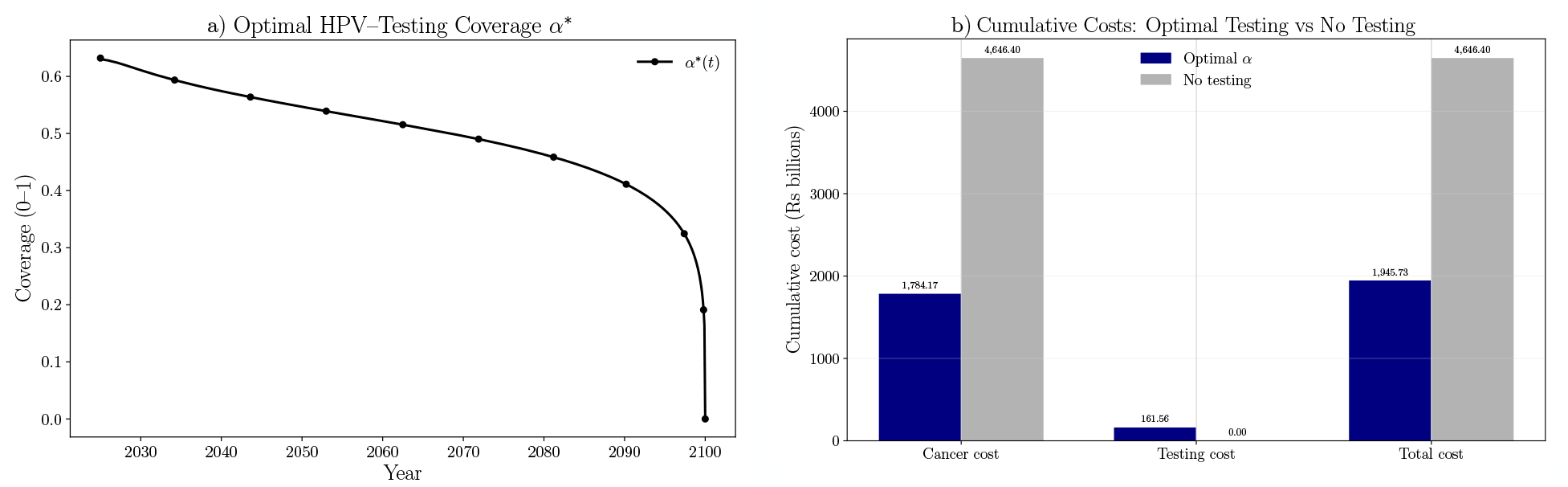
**Panel (a):** Optimal time-varying HPV-testing coverage *α*^*^(*t*) beginning in 2025. **Panel (b):** Cumulative cervical-cancer treatment cost, HPV-testing cost, and their sum (2025–2100) under the optimal policy (navy) versus a no-testing baseline (grey), highlighting the economic impact achieved by the optimal policy.

**Figure 8:**
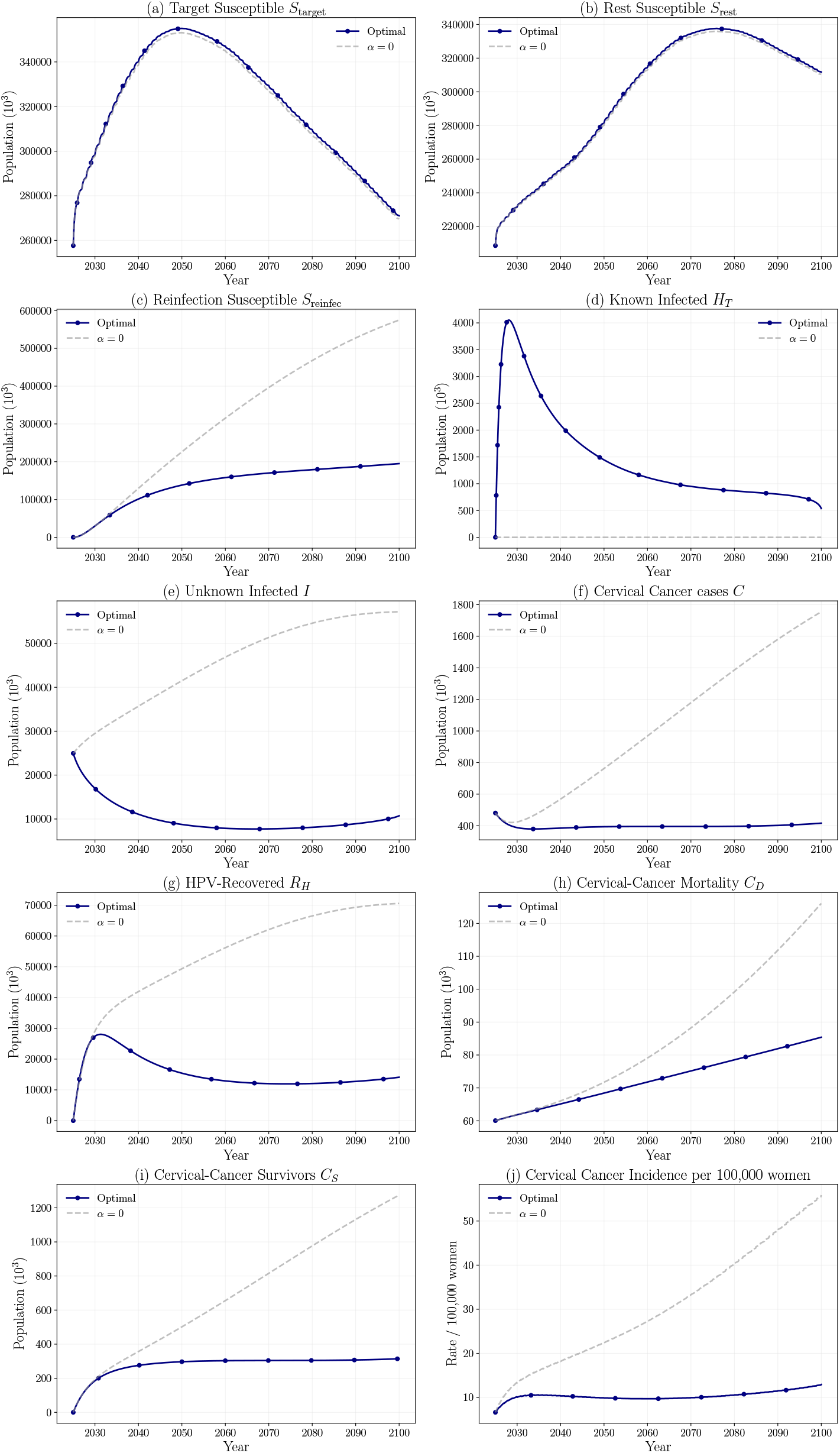
Dynamic HPV-testing policy obtained from optimal-control analysis (solid navy) versus the natural-history baseline with no testing (grey dashed), simulated from 2025 to 2100. Screening is restricted to women aged 30–60, and the coverage *α*^*^(*t*) is chosen to minimise the weighted sum of testing effort and cervical-cancer treatment costs. Panels (a)-(i) show the trajectories for nine state variables, and panel (j) shows annual cervical-cancer incidence per 100 000 women.

## 4. Discussion

### 4.1. Role of HPV testing in the reduction of HPV prevalence and cervical cancer HPV testing in creating awareness and behavioral changes

Mathematical models serve as powerful tools for predicting disease burden and evaluating intervention outcomes. This study integrates HPV testing as a distinct feature within a compartmental modeling framework to improve the estimation of HPV infection dynamics and cervical cancer burden, thus underscoring its role in reducing transmission and cancer incidence in the Indian context. The model demonstrated a strong correlation between the simulated outcomes and observed epidemiological patterns in cancer prevalence dynamics, as illustrated by the goodness of fit of the model (Figure 2). This is indicative of the model’s capability in accurately representing the complex epidemiological landscape of India for providing a robust foundation for policy-level forecasting. Furthermore, the forward simulations focusing on varying HPV testing coverage rates and target age groups demonstrate the potential impact of introducing HPV testing at the population level. For instance, higher testing rates led to increased identification of HPV infected individuals, enabling timely interventions (treatment) and thereby preventing the progression to precancerous and cancerous stages (Figure 4(j)). Similarly, expanding the target age group for HPV testing can augment early disease detection and limit infection transmission, subsequently reducing long-term cervical cancer burden (Figure 5(j)). The simulation studies on four states (refer to Figure 6) with different Human Development Index (HDI) and geographical locations indicate that the active HPV testing helps in the reduction of cervical cancer incidence over time. However, the long-term impact of the active HPV testing depends on the individual state in India. Our study on the cost-effective HPV testing intervention indicates the reduction in overall HPV infected cases, cervical cancer cases, improvement in the HPV-recovery, and constant cervical cancer incidence. It should be noted that the cost-effective HPV testing optimizes the trade-off between the cost of cancer treatment and the cost of active HPV testing. Hence, it can be seen from Figure 7(b) that the optimal HPV testing helps in reducing the total cost of cervical cancer burden by around 2.5 in the next 80 years.

Overall, the active HPV testing approach can serve as a vital complementary strategy, particularly in India, where the large target population size poses challenges in achieving prompt vaccine-induced herd immunity. Predicting baseline HPV prevalence through active testing can also prove beneficial in estimating cervical cancer reduction patterns not yet addressed by vaccination. It can be concluded that the integration of HPV testing in public health programs has the potential to induce behavioral changes at the individual as well as the community level. The HPV testing, coupled with the awareness about HPV infection, can promote informed health behavior and reduce stigma associated with STIs, ultimately generating greater acceptance of preventive interventions like vaccinations and early treatment, and will reduce cervical cancer incidence in India in the coming years.

## 5. Conclusions

In this work, we have developed a compartmental model that captures the dynamics of HPV infections and cervical cancer progression in the Indian context. By introducing an HPV testing infected compartment that enables age-specific testing, our model explains the impact of targeted HPV testing in reducing the incidence of infection, cancer prevalence, and related mortality. Numerical simulations highlight the benefits of testing in lowering both infection and cancer trajectories, with the impact varying with varying screening rates. The application of optimal control theory demonstrates that a properly chosen HPV testing strategy can offer a cost-effective balance between resource utilization for Testing and the healthcare costs related to cancer treatment.

Overall, these findings highlight the importance of policy-driven interventions, particularly in low-resource settings. Timely and appropriately scaled HPV screening programs, combined with effective public health initiatives and vaccination efforts, have the potential to substantially reduce HPV infections and the burden of cervical cancer.

## Data Availability

All data produced in the present work are contained in the manuscript

## Appendices

### A. Recruitment Rates (Λ ⋅ *p*) and Mortality Rates (*P*) as a function of time

Constant recruitment rates and mortality rates inadequately represent the dynamics of India’s population growth, as they result in a monotonic increase in population, contrary to projections. According to Probabilistic Population Projections from United Nations United Nations Department of Economic and Social Affairs, Population Division [31] Indian population is anticipated to peak at 2050, after which it is expected to stabilize. By modeling recruitment rates and mortality rates as functions of time, the population dynamics can be captured more accurately, reflecting these projections. Using the time-Series data for age-band wise population for females, for the years of **1950–2023** and using the population projections median data from [31], for the years of 2023-2100, we are modeling the age-wise population *Inflow* (Recruitment Rates): Inflow(*t*) and *Outflow* functions (Mortality Rates): Outflow(*t*), for the Target and Rest Age Bands, with their respective Λ, *p* and *P* functions as follows:

#### Discrete Data

From census (or UN/WPP) tables, we have discrete data on the annual series

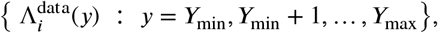

where 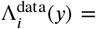 (in thousands) during calendar year *y* (births, ageing-in, net migration^*i*^, etc.).

#### Continuous-time model

The ODE system is written for continuous time *t* ∈ ℝ, by defining a *piece-wise/step-wise constant* reconstruction

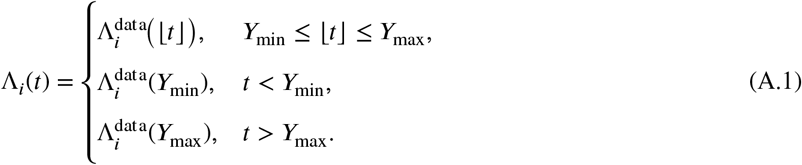

where * ⌊*t*⌋ picks the current calendar year (e.g. *t* = 2013 7 ↦ 2013). Equation A.1 states that Λ*_i_* (*t*) is *constant inside each year-interval* [*y, y*+1) and jumps only at integer years. These functions are visualized in Figure 9.

**Figure 9:**
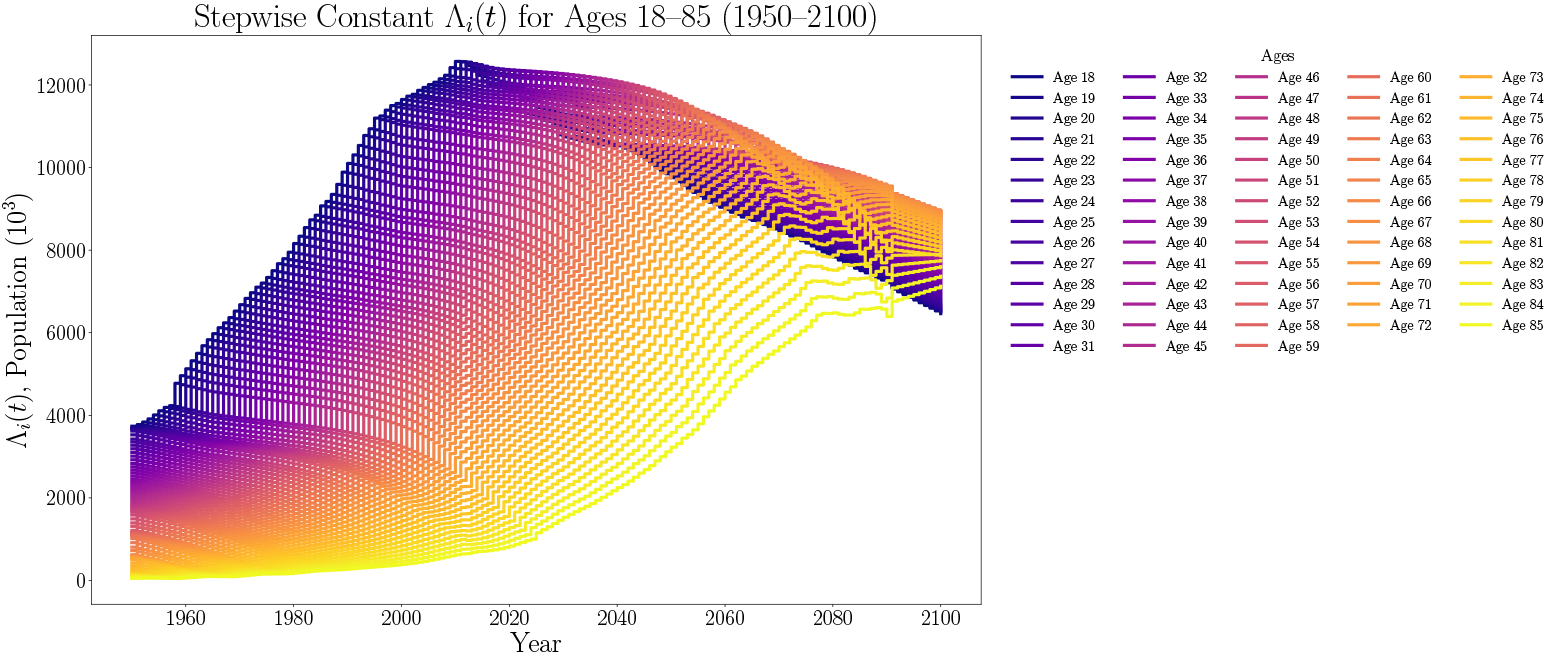
Piecewise/Step-wise constant constant functions Λ_*i*_(*t*) for females of ages *i* = 18, …, 85 in India (1950-2100)

Let the Target Age band for testing be defined as,

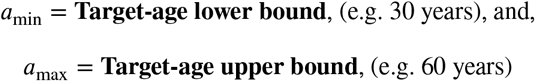

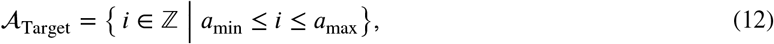

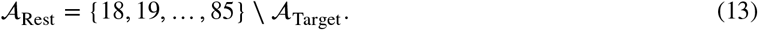

*Step-wise Constant Population functions*, Λ_*Target, Rest*_(*t*)

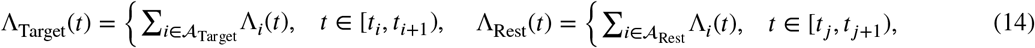

*Scaling Factors, p*_*Target, Rest*_(*t*)

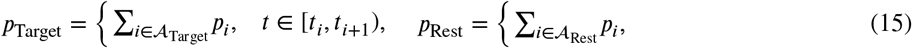

*Mortality/Outflow rates, P*_*Target, Rest*_(*t*)

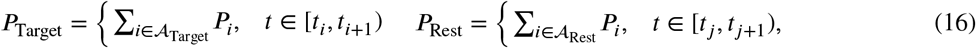

where *t*_*i*_ and *t*_*j*_ represent the start of each year from 1950 to 2100, with *i, j* ∈ {1950, 1951, …, 2100}. *p*_*i*_ & *P*_*i*_ ∀*i* ∈ {18, …, 85}, were obtained by Least-Squares fitting to the original Indian population dynamics.

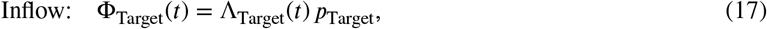

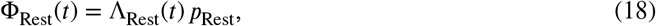

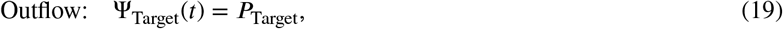

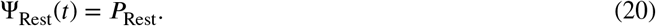

where Inflow represents Total entry rate (births + ageing-in ± migration), in thousands and Outflow represents the Total exit rate (natural mortality, migration, ageing-out) in thousands, respectively, for both Target and Rest Age Bands

#### B. Comparison of Detection of HPV Infected Females by Different HPV Screening Methods Based on Their Sensitivity

Figure 10 shows the isolated dynamics of known infected state four commonly used screening tests: Pap cytology, visual inspection with acetic acid (VIA), liquid-based cytology (LBC) and high-risk HPV DNA testing—ranked by their analytical sensitivities (*H* ^+^ = 0.70, 0.75, 0.85, and 0.95, respectively) with the screening programs starting in 2025, with a Screening Coverage rate of 90% and with the Target Age band for screening being females aged between 30-60. The trajectories for the *H*_*T*_ state show how many women enter the *HPV known infected* compartment (*H*_*T*_) once universal screening is introduced. Because HPV DNA testing detects viral nucleic acid directly, its high sensitivity captures almost the entire pool of latent and low-grade infections, pushing the *H*_*T*_ curve well above those for VIA, Pap, and LBC.

**Figure 10:**
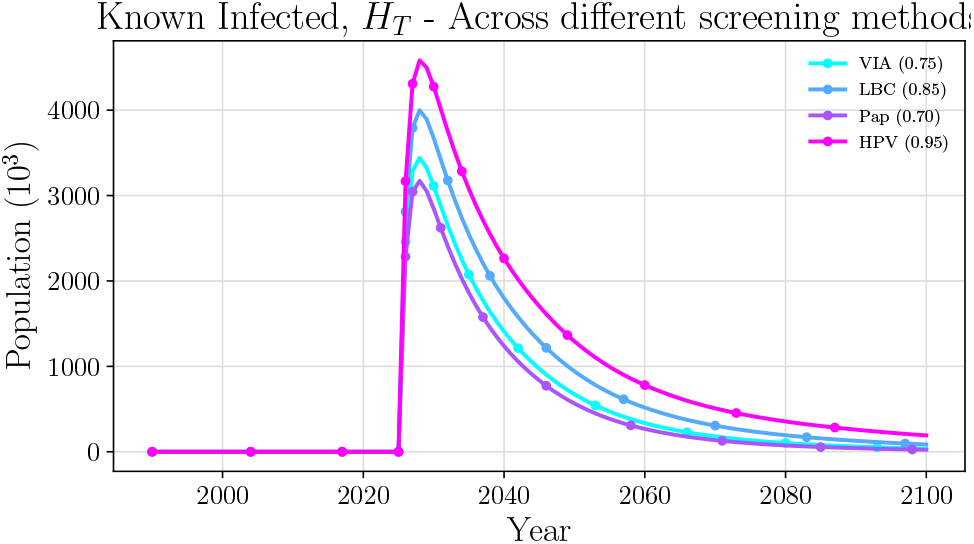
Modelled number of HPV-tested positive Infected women (*H*_*T*_) under four screening tests with increasing test sensitivity *H* ^+^: Pap cytology (0.70), VIA (0.75), LBC (0.85) and HPV DNA testing (0.95). Higher sensitivity translates into substantially more infections detected.

#### C. Basic Reproduction Number ℛ_*o*_

The basic reproduction number (denoted by ℛ_*o*_) is the average number of secondary infections caused by a single infectious individual in a fully susceptible population during its entire cycle of infectiousness (Diekmann et al. [8]). We calculate the ℛ_*o*_ when *α* = 0, and the model follows **SICR** path. Using the method of next generation matrix developed by Van den Driessche and Watmough [9], let *X* = (*I, C, S*_target_, *S*_rest_, *S*_*R*_, *R*_*H*_), then system can be written as

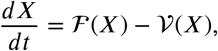

where ℱ (*X*) contains terms that represent new infections and 𝒱 (*X*) contains all other transitions out of infected compartments. For *ℱ* (*X*), only the *I* compartment contributes to the new infections.

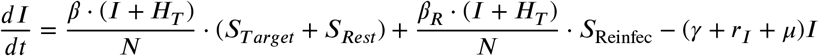

At Disease Free Equilibrium (DFE), *I* = *C* = *R*_*H*_ = *H*_*T*_ = 0 and the susceptible stocks are 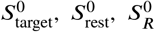 with 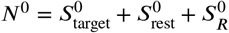. Hence at DFE, we have,

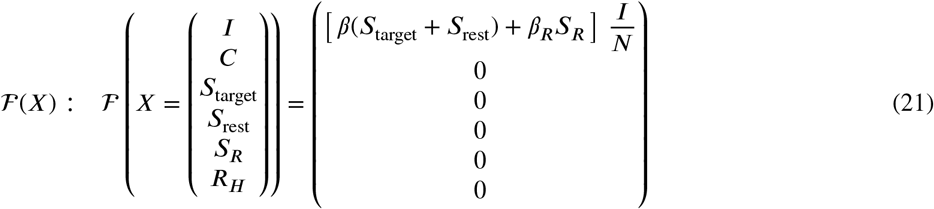

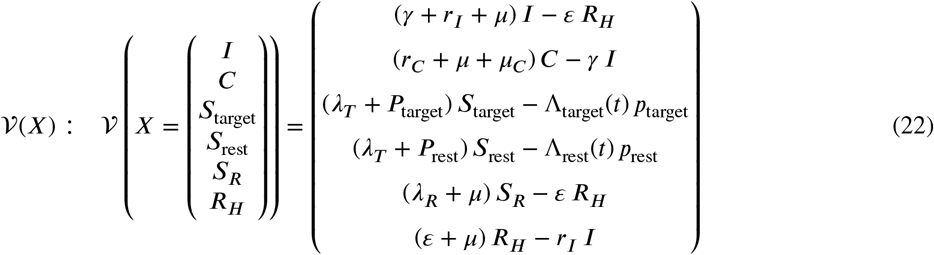

where 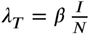 and 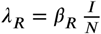. Linearizing around Disease Free Equilibrium, 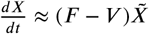,

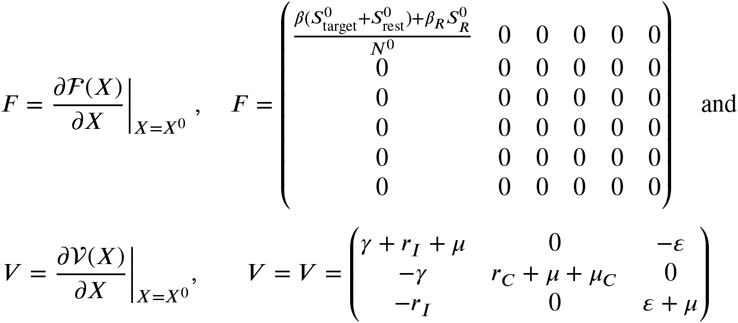

The next generation matrix is ℱ𝒱^−1^, then the Reproduction number ℛ_*o*_ is

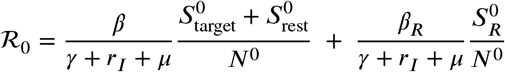

The stability of the DFE depends on the basic reproduction number: If ℛ_0_ *<* 1: The DFE is stable (the disease dies out). If ℛ_0_ *>* 1: The DFE is unstable (the disease spreads). Since our ℛ_0_ *>* 1, the DFE is unstable.

#### D. Sensitivity Analysis of Basic Reproduction Number, ℛ_0_

The Sensitivity indices of the reproduction number are computed to identify parameters with large impact on ℛ_0_. To analyze the sensitivity of the basic reproduction number (ℛ_0_) with respect to key model parameters, we begin by considering its analytical expression. With *α* = 0 and following the SICR disease progression pathway, ℛ_0_ is given by:

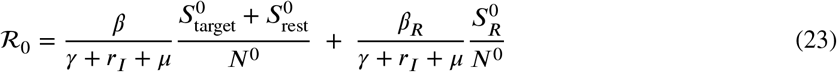

To quantify the relative influence of each parameter *p* on ℛ_0_, we compute the **Normalized Sensitivity Index**, This index measures the relative change in ℛ_0_ due to a relative change in *p*.

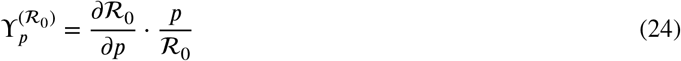

We derive the partial derivatives of ℛ_0_ with respect to each parameter in the second column of Table 2. Then by substituting baseline parameter values, we compute the sensitivity indices numerically (fourth column of Table 2). This allows us to determine the most influential parameters affecting ℛ_0_. Figure 11 describes the the effect of different parameters on ℛ_0_.

**Table 2.** Sensitivity analysis of ℛ_0_ for the HPV–testing model (baseline ℛ_0_ = 1.1026)

| Parameter $p$ | $\frac{\partial \mathcal{R}_0}{\partial p}$ | Normalised index $\Upsilon_p^{(\mathcal{R}_0)}$ | Evaluated at baseline |
| --- | --- | --- | --- |
| $\beta$ (Primary Transmission rate) | $\frac{f_T}{\gamma + r_I + \mu}$ | $\frac{\beta f_T}{\beta f_T + \beta_R f_R}$ | +0.974 |
| $\beta_R$ (Reinfection Transmission rate) | $\frac{f_R}{\gamma + r_I + \mu}$ | $\frac{\beta_R f_R}{\beta f_T + \beta_R f_R}$ | +0.027 |
| $\gamma$ (Progression to Cancer) | $-\frac{\beta f_T + \beta_R f_R}{(\gamma + r_I + \mu)^2}$ | $-\frac{\gamma}{\gamma + r_I + \mu}$ | -0.003 |
| $r_I$ (HPV Recovery rate) | $-\frac{\beta f_T + \beta_R f_R}{(\gamma + r_I + \mu)^2}$ | $-\frac{r_I}{\gamma + r_I + \mu}$ | -0.967 |
| $\mu$ (Natural mortality) | $-\frac{\beta f_T + \beta_R f_R}{(\gamma + r_I + \mu)^2}$ | $-\frac{\mu}{\gamma + r_I + \mu}$ | -0.030 |

**Figure 11:**
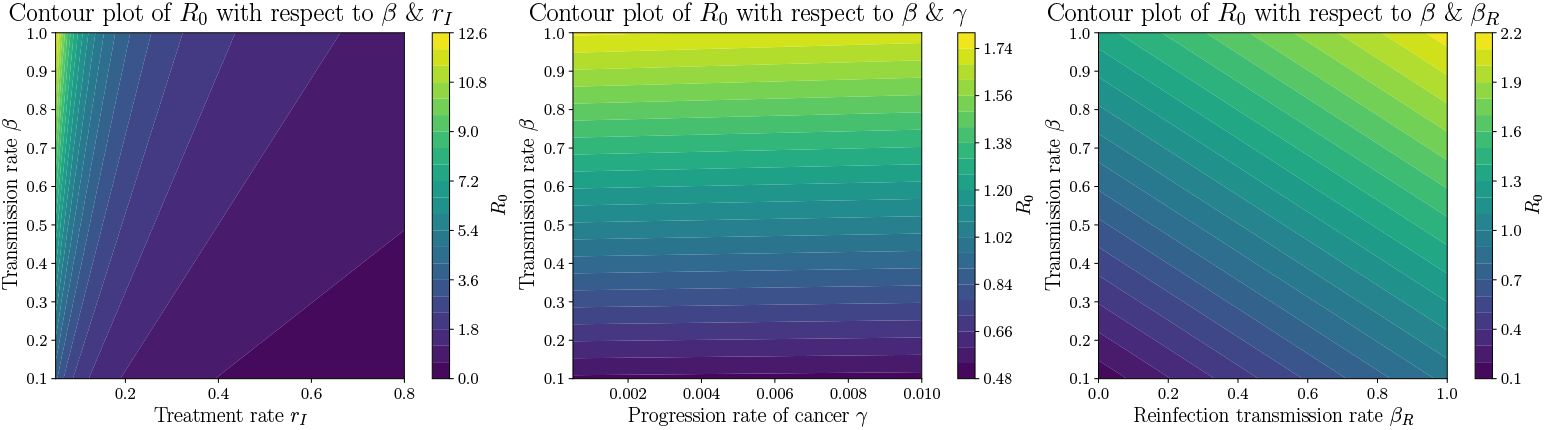
Contour plots of ℛ_0_ with different parameter combinations

#### E. Monte Carlo Parameter Uncertainty Analysis

To assess the sensitivity of our mathematical model to variability in key epidemiological parameters, we performed a Monte Carlo simulation using 1000 randomly sampled parameter sets. The ranges for the parameters represent either literature-reported confidence intervals or intervals obtained from previous parameter fitting simulations. The parameters considered in this analysis include *H* ^+^, *β, β*_*R*_ *μ, γ, r*_*I*_, *r*_*C*_, *ε, μ*_*C*_, *ω* and *r*_*H*_. Figure 12 shows the output of this Monte Carlo Parameter Sampling Analysis for the prevalence and incidence of HPV infection (compartment *I*) and cancer (compartment *C*), which are expressed per 100,000 women. At each time point, the 2.5th, 50th (median), and 97.5th percentiles were computed over the 1000 simulations. These percentiles are visualized in Figure 12 as shaded regions (from 2.5th to 97.5th percentile) with the median curve overlaid. For comparison, the simulation result using the original parameter set is also plotted. Table 3 summarizes the updated Monte Carlo parameter statistics, including the original values and the corresponding median as well as the 2.5th to 97.5th percentile intervals.

**Table 3.** Monte Carlo Parameter Statistics and Original Values.

| Parameter | Original Value | Median | 2.5th – 97.5th Percentile |
| --- | --- | --- | --- |
| $H^+$ | 0.9500 | 0.9036 | [0.8039, 0.9945] |
| $\beta$ | 0.5433 | 0.5446 | [0.4545, 0.6448] |
| $\beta_R$ | 0.4932 | 0.5072 | [0.4045, 0.5956] |
| $\mu$ | 0.0139 | 0.0127 | [0.0102, 0.0149] |
| $\gamma$ | 0.0013 | 0.0055 | [0.0013, 0.0097] |
| $\theta$ | 0.0001 | 0.0006 | [0.0001, 0.0010] |
| $r^H$ | 0.9000 | 0.6997 | [0.5099, 0.8899] |
| $r^I$ | 0.4500 | 0.5728 | [0.4555, 0.6938] |
| $r^C$ | 0.1340 | 0.1296 | [0.1009, 0.1584] |
| $\varepsilon$ | 0.3500 | 0.2971 | [0.0824, 0.5300] |
| $\mu^C$ | 0.0009 | 0.0005 | [0.0001, 0.0010] |
| $\omega$ | 0.1600 | 0.1595 | [0.1504, 0.1694] |

**Table 4.**
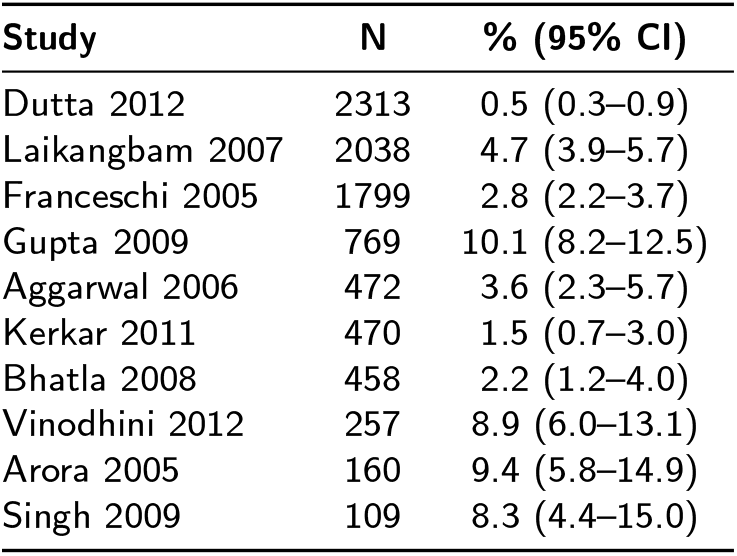
HPV 16 prevalence among women with normal cervical cytology in India, by study (HPV Information Center [4])

| Study | N | % (95% CI) |
| --- | --- | --- |
| Dutta 2012 | 2313 | 0.5 (0.3–0.9) |
| Laikangbam 2007 | 2038 | 4.7 (3.9–5.7) |
| Franceschi 2005 | 1799 | 2.8 (2.2–3.7) |
| Gupta 2009 | 769 | 10.1 (8.2–12.5) |
| Aggarwal 2006 | 472 | 3.6 (2.3–5.7) |
| Kerkar 2011 | 470 | 1.5 (0.7–3.0) |
| Bhatla 2008 | 458 | 2.2 (1.2–4.0) |
| Vinodhini 2012 | 257 | 8.9 (6.0–13.1) |
| Arora 2005 | 160 | 9.4 (5.8–14.9) |
| Singh 2009 | 109 | 8.3 (4.4–15.0) |

**Figure 12:**
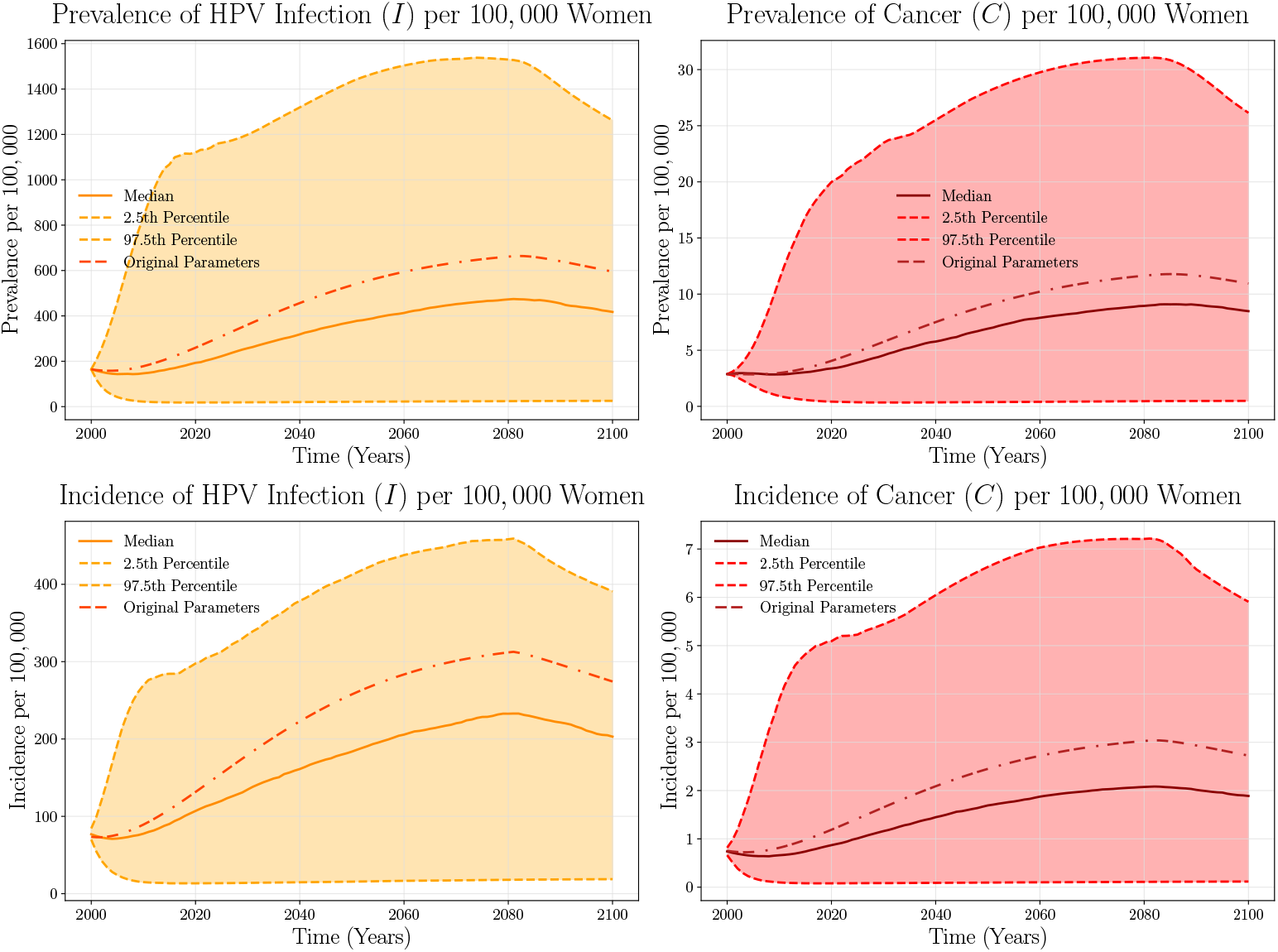
Monte Carlo Parameter Sampling Analysis. Shaded regions denote the 2.5th to 97.5th percentile bounds across 1000 simulations. The solid curves represent the median projections, and the red curves correspond to the simulation using the original parameter set. The left panels (orange shades) pertain to HPV infection outcomes, while the right panels (red shades) pertain to cancer outcomes.

#### F. Necessary Condition of Optimality, Algorithms and Implementations for Optimal Control Problem 11

##### Partial derivatives required by Pontryagin’s principle

###### Running-cost gradients

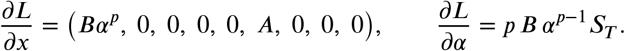

*Useful derivatives of the force of infection*. Let *Z* = *I* + *H*_*T*_. Then

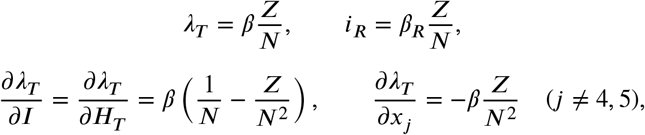

and identical formulas with *β* ↦ *β*_*R*_ for *i*_*R*_.

*Gradient of H w*.*r*.*t. α*. Only *f*_1_, *f*_4_, *f*_5_ depend on *α*:

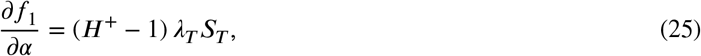

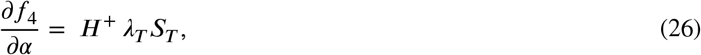

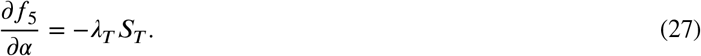

Hence

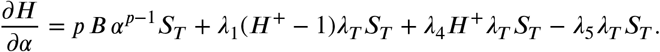

**Optimality condition** Setting *∂H* /*∂α* = 0 and dividing by *S*_*T*_ *>* 0 gives

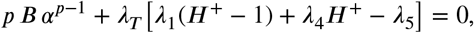

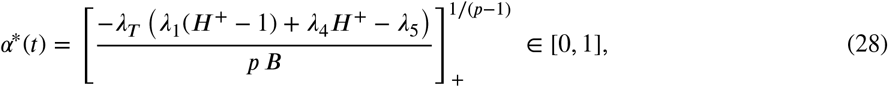

The detailed implementation of Optimal Control for HPV testing with its respective adjoint (costate) system (backwards equations) and the pseudo-code for the Numerical Solution for the implementation of Optimal Control using forward-backward sweep with Runge Kutta 4 method are given as follows.

##### Adjoint (costate) system (backward equations)

Pontryagin’s principle demands _*i*_ = −*∂H* /*∂x*_*i*_.

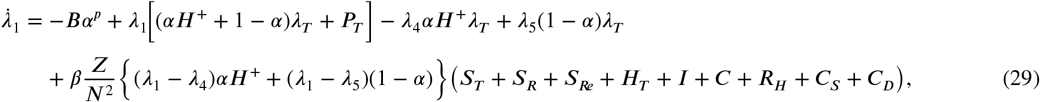

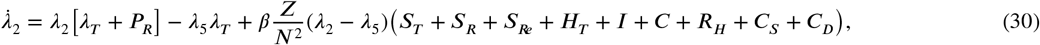

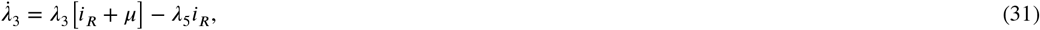

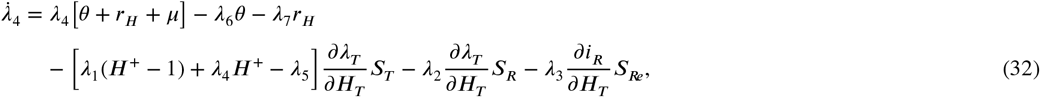

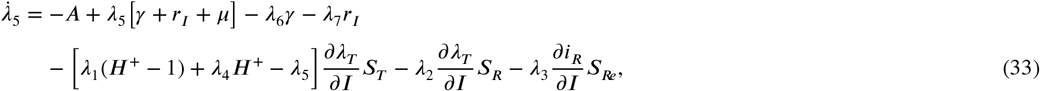

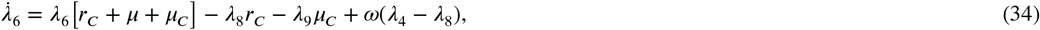

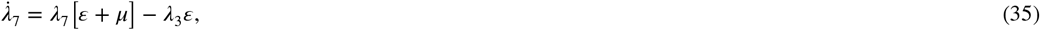

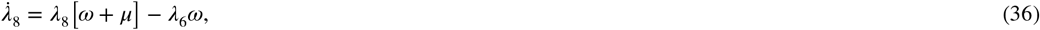

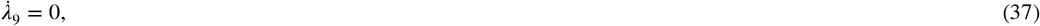

with terminal conditions *λ*_*i*_(*t*_*f*_) = 0. The pseudo Code for Numerical solution using the forward–backward sweep with the runge kutta 4th order formula is provided in Algorithm 1.

#### G. Rationale for the Cervical-Cancer Treatment Cost and HPV Testing Cost Used in the Budget Constrained Simulation

A detailed mixed micro-costing study by Singh et al. [29], conducted in the 2016–17 price year at the Post-Graduate Institute of Medical Education and Research (PGIMER), Chandigarh, followed 248 cervical-cancer patients and computed a *Health Benefit Package* (HBP) cost for each primary treatment arm by summing provider-side economic costs and mean patient out-of-pocket expenditure (OOPE). The study reported an HBP of Rs. 45 364 (≈ US$544) for interstitial brachytherapy, Rs. 56,538 (≈ US$679) for radical hysterectomy, and Rs. 60,422 (≈ US$725) for a complete course of 3-D conformal external-beam radiotherapy. Because the radiotherapy pathway represents the costliest standard regimen and therefore sets a prudent upper bound within the public sector, the present simulation adopts a rounded per-patient treatment cost of **Rs 60,000 (**≈ **US$720)**, providing a conservative yet empirically grounded estimate for budget-impact calculations. Market quotations from leading private diagnostics chains in 2025 place the price of a single high-risk HPV DNA test between Rs. 1,450 and Rs.3,075. Allowing an additional margin for specimen-collection logistics, consumables, and regional price variation, and noting that historical trial-based estimates uprated to 2025 Values still fall below this bracket—this study adopts a conservative upper-bound cost of **Rs. 3,000 (**≈ **USD 36)** per woman screened. Using this worst-case figure avoids underestimating programme expenditure.

##### Algorithm 1

Optimal HPV–testing policy via forward–backward sweep (RK4)

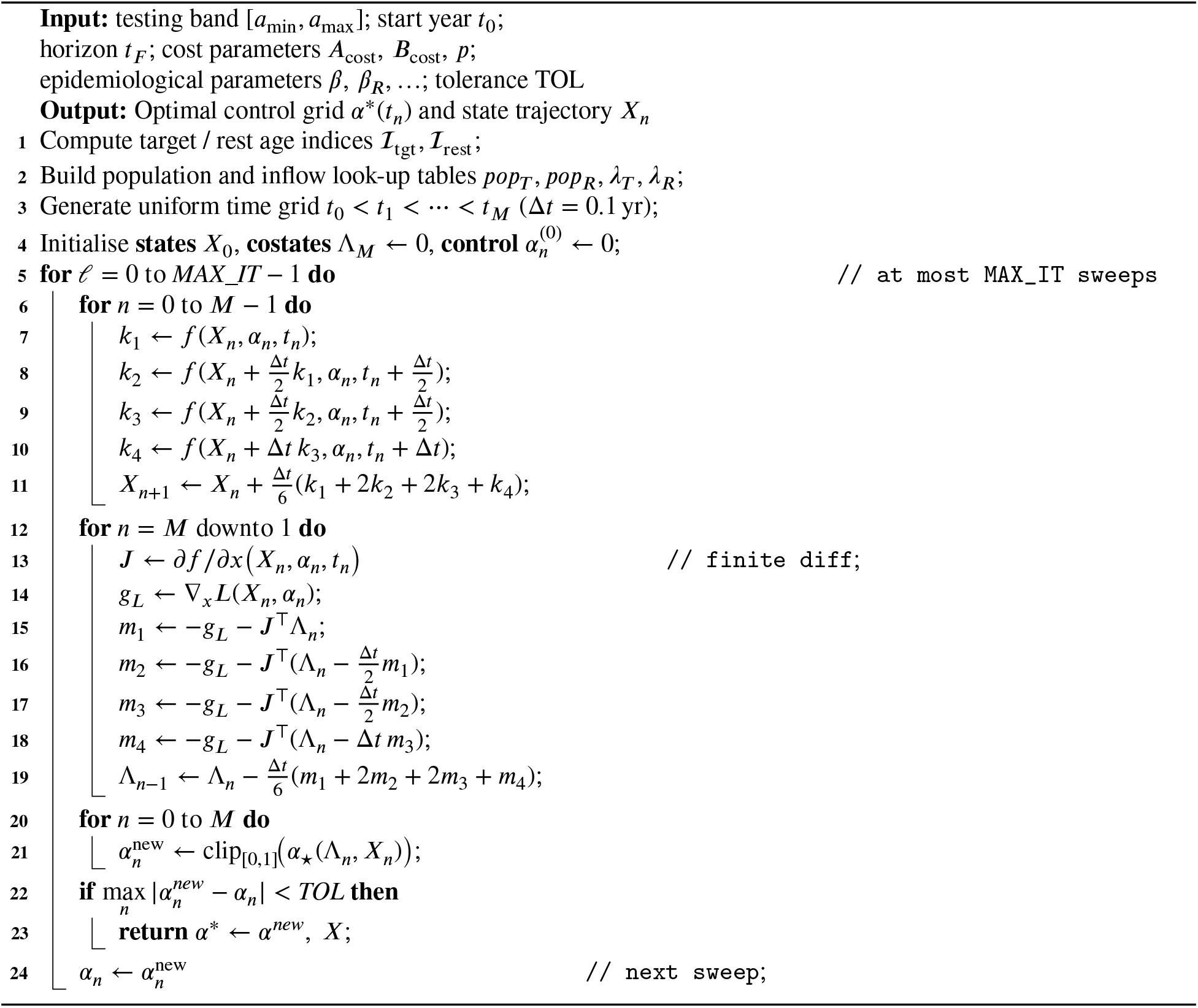

## Acknowledgment

Funding for this study was provided by the Gates Foundation (NV-044445).

## Disclaimer

This work/opinion is based on research findings by the authors and not the opinion of the government.

## Credits

**Arvinth S**: Data curation, Formal analysis, Investigation, Methodology, Validation, Visualization, Writing – original draft, Writing – review & editing. **Rashmi Tiwari**: Conceptualization, Data curation, Formal analysis, Investigation, Methodology, Validation, Writing – original draft, Writing – review & editing. **Vishnu Tej**: Data curation, Formal analysis, Investigation, Methodology. **Piyush Sawarkar**: Data curation, Formal analysis, Investigation, Methodology. **Prerna Bhalla**: Data curation, Formal analysis, Investigation, Methodology, Validation, Visualization, Writing – review & editing. **Shekar Sivasubramanian**: Formal analysis, Investigation, Methodology. **Ajit Rajwade**: Conceptualization, Formal analysis, Funding acquisition, Investigation, Methodology, Project administration, Resources, Supervision, Writing – review & editing. **Ganesh Ramakrishnan**: Conceptualization, Formal analysis, Funding acquisition, Investigation, Methodology, Project administration, Resources, Supervision, Writing – original draft, Writing – review & editing. **Nirav Bhatt**: Conceptualization, Formal analysis, Funding acquisition, Investigation, Methodology, Project administration, Resources, Supervision, Writing – original draft and Writing – review & editing.

## Footnotes

1 USD conversions use the mid-2025 exchange rate of 1 INR 0.012 USD.

